# Evaluating the comparability of osteoporosis treatments using propensity score and negative control outcome methods in UK and Denmark electronic health record databases

**DOI:** 10.1101/2023.10.02.23296212

**Authors:** Trishna Rathod-Mistry, Eng Hooi Tan, Victoria Y Strauss, James O’Kelly, Francesco Giorgianni, Richard Baxter, Vanessa C Brunetti, Alma Becic Pedersen, Vera Ehrenstein, Daniel Prieto-Alhambra

**Author notes:** Corresponding author: Daniel Prieto-Alhambra, Address: Botnar Research Centre Nuffield Department of Orthopaedics, Rheumatology, and Musculoskeletal Sciences, University of Oxford, OX3 7LD, UK. Joint first author (Trishna Rathod-Mistry, Eng Hooi Tan).

## Abstract

Evidence on the comparative effectiveness of osteoporosis treatments is heterogeneous. This may be attributed to different populations and clinical practice, but also to differing methodologies ensuring comparability of treatment groups before treatment effect estimation and the amount of residual confounding by indication. This study assessed the comparability of denosumab vs oral bisphosphonate (OBP) groups using propensity score (PS) methods and negative control outcome (NCO) analysis. A total of 280,288 women aged ≥50 years initiating denosumab or OBP in 2011-2018 were included from the UK Clinical Practice Research Datalink (CPRD) and the Danish National Registries (DNR). Balance of observed covariates was assessed using absolute standardised mean difference (ASMD) before and after PS weighting, matching, and stratification, with ASMD >0.1 indicating imbalance. Residual confounding was assessed using NCOs with ≥100 events. Hazard ratio (HR) and 95% confidence interval (CI) between treatment and NCO was estimated using Cox models. Presence of residual confounding was evaluated with two approaches: (1) >5% of NCOs with 95% CI excluding 1, (2) >5% of NCOs with an upper CI <0.75 or lower CI >1.3. The number of imbalanced covariates before adjustment (CPRD 22/87; DNR 18/83) decreased, with 2-11% imbalance remaining after weighting, matching or stratification. Using approach 1, residual confounding was present for all PS methods in both databases (≥8% of NCOs). Using approach 2, residual confounding was present in CPRD with PS matching (5.3%) and stratification (6.4%), but not with weighting (4.3%). Within DNR, no NCOs had HR estimates with upper or lower CI limits beyond the specified bounds indicating residual confounding for any PS method. Achievement of covariate balance and determination of residual bias were dependent upon several factors including the population under study, PS method, prevalence of NCO, and the threshold indicating residual confounding.

## Introduction

Routinely collected data from clinical practice settings have been used to evaluate the real-world effectiveness of osteoporosis treatments in reducing risk of fracture amongst postmenopausal women. Choice of treatment is dependent on a range of factors including patient medical history such as previous fractures, falls and treatment, patient and clinician treatment preference, and effectiveness of treatment for specific fracture sites (1). According to the European guidance and clinical guidelines in the UK, first line treatment typically includes oral bisphosphonates (alendronate, risedronate, ibandronate) and second line includes denosumab if bisphosphonates are not suitable or tolerated (1–4).

Real-world evidence on the effectiveness of denosumab vs. oral bisphosphonates on fracture risk is inconsistent (5). Studies have shown denosumab was equally effective as oral bisphosphonates in reducing the risk of non-vertebral (6) and hip (7) fractures using US claims and Danish registry data, respectively. However, an analysis of Spanish pharmacy data showed that denosumab reduced the risk of hip and any type of fracture more than oral bisphosphonates (8). Although these differing results may reflect genuine differences in effectiveness due to study populations, settings, treatment guidelines, and comparator groups, confounding by indication may also explain these results, due to the inherent differences between patients using first and second-line treatments. For instance, patient characteristics are likely to differ between treatment groups, which may affect fracture prognosis.

To account for measured confounding, propensity score (PS) methodology can be used to create balanced treatment groups with respect to measured covariates, such as age and history of fracture, by using the PS in matching, stratification, or inverse probability treatment weighting (IPTW) (9, 10). However, information on important confounders such as bone mineral density, may be missing in administrative databases, and thus cannot be adjusted for using PS methods, which can lead to unmeasured confounding.

Negative control outcomes (NCOs) can be used to minimize residual confounding by unmeasured covariates. NCOs are outcomes known not to be causally associated with treatment. NCO methods have been developed to detect the presence of unmeasured (residual) confounding, when an association is found between the treatment and NCO (11, 12). However, there are currently no gold standards or guidelines on the threshold for comparability between treatment groups using NCO. Previous studies have defined bias as non-null effect of an NCO using risk difference and confidence interval (CI) (13, 14), as well as more than 5% of a large set of NCOs having 95% CI excluding 1 (15).

This study aims to determine whether cohorts who received denosumab or oral bisphosphonates were comparable using PS matching, stratification and IPTW, and by applying different rules for presence of residual confounding via NCO analysis, in two European databases.

The objectives were to:

1. Describe osteoporosis treatment groups, denosumab vs oral bisphosphonates, with respect to demographics, clinical history, and prior medication use
2. Assess whether treatment groups are comparable on measured covariates after PS matching, stratification and IPTW.
3. Detect the presence of residual confounding using different stringency rules via NCO analysis within each PS method.
4. Assess whether comparability is achieved in subgroups of older patients, post fracture patients, and patients with potentially three years of follow-up, plus a post-hoc analysis of second line users.

## Methods

### Study design and setting

A retrospective, new user and new switcher, active comparator cohort study was implemented (16).

We used data from two European countries, including the UK Clinical Practice Research Datalink (CPRD) GOLD (17) and AURUM (18), which are primary care databases of medical records from general practitioners. CPRD was linked to the following databases: Hospital Episode Statistics Admitted Patient Care, Office for National Statistics mortality data, and the Index of Multiple Deprivation (IMD). Practices that appeared in both the GOLD and AURUM databases were retained in AURUM and excluded from GOLD.

We also used data from the Danish National Registries (DNR), which contains linked data from the Danish Civil Registration System (19), the Danish National Patient Registry (20), and the Danish National Prescription Registry (21). DNR contains dispensations in outpatient pharmacies, inpatient and outpatient hospital clinics encounters, and complete follow-up until death or emigration, set within the universal Danish healthcare system (22).

### Participants

We included women aged ≥50 years at the time of initiating denosumab or oral bisphosphonates between 01 January 2011 to 31 December 2018 for UK patients and 01 January 2011 to 31 December 2017 for Danish patients. For the new user cohort, the date of initiating denosumab or bisphosphonates was defined as the index date (Figure 1). We included patients who had no history of these treatments in the year before the index date prior to treatment initiation and were registered with their practice for at least one year before index.

**Figure 1:**
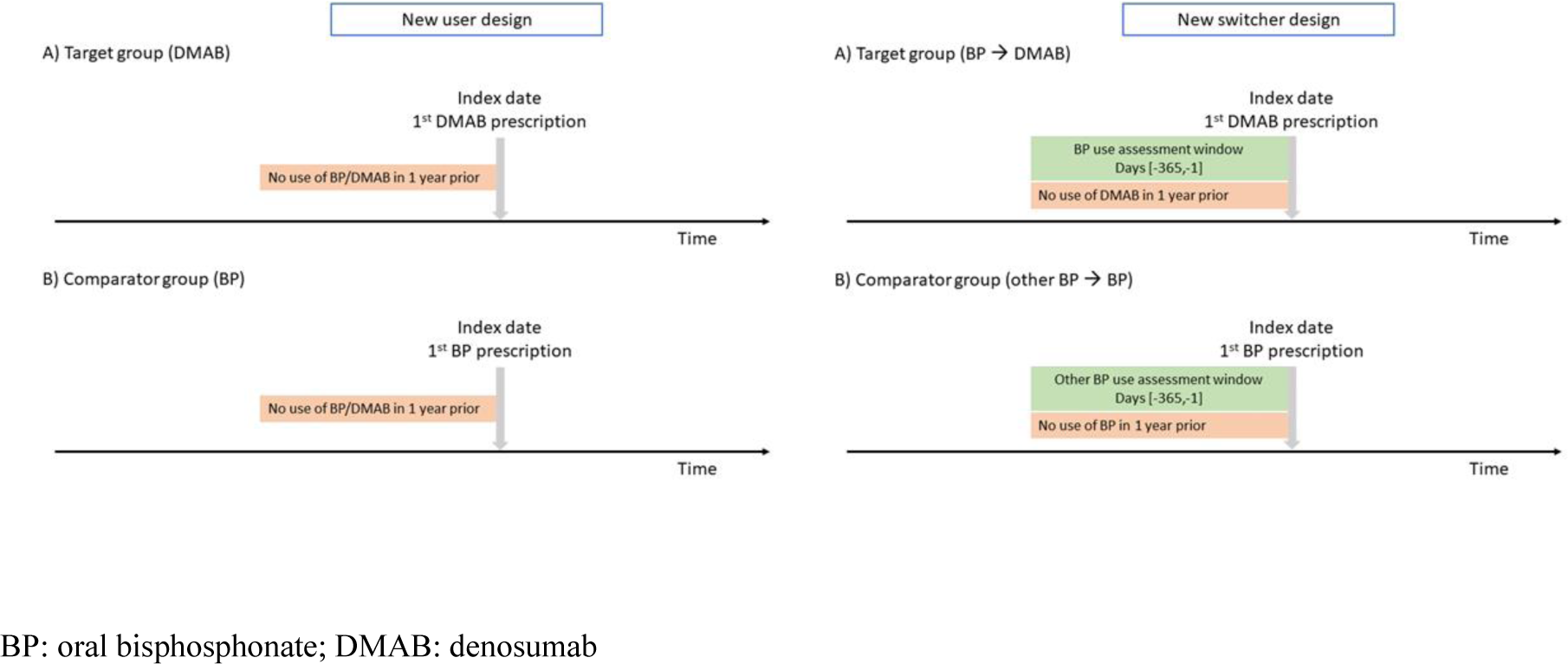
New user and new switcher study designs.

To reflect the treatment guidelines in place in the UK and Denmark, where oral BPs are recommended as first-line treatments, and denosumab is recommended as second-line treatment, we assembled a cohort of recent treatment switchers in post-hoc analyses. This approach allowed us to compare patients at a similar stage of treatment. The treatment groups were re-defined as follows, (1) denosumab users included patients who switched from bisphosphonates to denosumab with no previous use of denosumab in the one year prior, and (2) bisphosphonate users included patients who switched from one oral bisphosphonate to a new oral bisphosphonate and had no previous prescription for the new bisphosphonate in the one year prior (Figure 1). The index date was defined as the date of the initiation of the new treatment.

Patients were excluded if they had any of the following diagnoses in the five years prior to the index date, or treatments in the year before the index date: Paget’s disease of bone, cancer (except for non-melanoma skin cancer) or its associated treatments (hormonal, endocrine, or radiation therapies), end stage renal disease, or prescription for both denosumab and oral bisphosphonates on the index date.

Patients were followed from the index date until the earliest censoring event occurrence: NCO (defined below), death, moved practice, end of study period (31^st^ December 2019 for CPRD and 31^st^ December 2018 for DNR), drug discontinuation or treatment group switch, diagnosis of cancer (except for non-melanoma skin cancer) or its associated treatments (hormonal, endocrine, or radiation therapies), end stage renal disease, or a maximum three-year follow-up.

### Treatment groups

The treatment groups were patients initiating (new user) or switching to (switcher) either (1) oral bisphosphonates (alendronate 10mg/day or 70mg/week, ibandronate 150mg/month, risedronate 5mg/day or 35mg/week) or (2) denosumab (60mg/6 months). We used an as-treated exposure definition, where patients were required to be on the initiated or new treatment throughout follow-up. Successive prescriptions were considered as a continuous treatment episode if there was no more than 90 days between prescriptions. Discontinuation was defined as a gap of more than 90 days between prescriptions; date of discontinuation was therefore defined as the end date of drug duration of the last prescription plus an additional 90 days (23).

For bisphosphonates, duration of prescription was calculated as the ratio of the number of tablets and daily dose; if duration was missing, the default of 30 days was used. For denosumab, duration was defined as a default of 180 days in line with the dosing interval indicated for osteoporosis. The end date of prescription was defined as the date of the prescription plus duration of prescription.

### Propensity score analysis and covariates

PS expressed the probability of being assigned denosumab rather than an oral bisphosphonate conditional on measured covariates. Logistic regression estimated the PS for each patient using over 80 covariates that are known to be associated with risk of fractures or falls (Table S1) (13). Covariates with more than 10 patients in each treatment group were included in the logistic regression model. The PS was then used in three ways to create comparable treatment groups.

**In PS matching**, each denosumab user was matched with up to five oral bisphosphonate users that had the closest PS within a calliper width of 0.2 of the pooled standard deviation of the logit of the PS (24, 25). In addition, exact matching was performed on year of index date as it was deemed to be an empiric confounder.

**In PS stratification**, all patients in the study population were divided into ten mutually exclusive strata based on the ranked PS distribution of the denosumab group. Within each stratum, the distribution of covariates between treatment groups should be similar.

**In IPTW**, each patient was assigned a weight equal to the inverse of their propensity score (1/PS), to create a pseudo-population with a balanced covariate distribution (26). Stabilised weights were calculated by multiplying the weights by the proportion of patients on each treatment to reduce large weights (27). To further minimise large weights, weights were then truncated at 99% percentile of distribution of the stabilised weights (26). Only the IPTW was used in the new switcher cohort.

Balance of each covariate between treatment groups was assessed using the absolute standardised mean difference (ASMD) before and after matching, stratification and IPTW. An ASMD less than 0.1 indicates the treatment groups were balanced for that covariate.

### Negative control outcome analysis

NCOs are defined as outcomes not causally associated with the treatment of interest, except through shared confounders (11, 12). If the estimated association between treatment and NCO is non-null, one can determine whether there is evidence of residual confounding suggesting treatment groups are not comparable with respect to unmeasured covariates.

A preliminary and non-exhaustive list of NCOs were identified, including fracture and non-fracture NCOs. Based on the FREEDOM trial (28), there was no effect of treatment on risk of fracture in the first three months; therefore, a fracture (hip, vertebral, radius, ulna, wrist, humerus, pelvis, or shoulder) occurring in the first three months of treatment was considered a NCO (29). In addition, NCOs were also sourced from previous studies (13, 30, 31), and from an automated method identifying potential NCOs (32). The maximum follow up for fracture NCOs was three months. The preliminary list of NCOs is listed in Table S1.

Analysis for a specific NCO was conducted if there were ≥100 events in total. The association between treatment and NCO was estimated using the Cox Proportional Hazards model with robust standard errors, adjusted for year of index and imbalanced covariates with ASMD ≥0.1 for each PS method (33). The estimated hazard ratio (HR) and 95% CI was used to assess for the presence of residual confounding based on two approaches:

**Approach 1:** There were more than 5% of NCOs having a 95% CI excluding 1; the denominator was the number of NCOs with ≥100 events and the numerator was the number NCOs with a CI excluding the null value of one.

**Approach 2:** There are more than 5% of NCOs having an upper CI of HR <0.75 or lower CI >1.30; the denominator was the number of NCOs with ≥100 events and the numerator was the number of NCOs having CI above or below the specified cut-off values.

Treatment groups were deemed comparable if residual confounding was absent in both Approach 1 and Approach 2.

### Subgroup analysis

Comparability was further assessed in three subgroups within the PS method that had achieved comparability. The three subgroups included older patients aged ≥65 years, post-fracture patients, and patients with potentially three years of follow-up. The PS was re-estimated for the patients eligible for analysis.

### Results

Analysis was performed separately for CPRD (combining the GOLD and AURUM databases) and DNR.

### CPRD

#### New user cohort

Of the 200,179 eligible patients, 6,528 were denosumab users and 194,191 were bisphosphonate users (Table 1). Most demographics, comorbidities, and prescriptions were balanced between denosumab and bisphosphonate users; however, there were 22 (25.3%) covariates whose distributions were imbalanced (Table 2). Denosumab users were observed to be older, had a higher number of GP visits, hospital admissions, fractures, longer duration of cumulative oral bisphosphate use, higher prevalence of calcium or vitamin D, proton pump inhibitor, lower prescriptions for non-steroidal anti-inflammatory drugs and corticosteroids than bisphosphonate users.

**Table 1:** Baseline patient characteristics of new users of denosumab and oral bisphosphonates.

|  | CPRD |  | DNR |  |
| --- | --- | --- | --- | --- |
| Characteristic | Denosumab<br>N = 6,528 | Oral<br>bisphosphonate<br>N = 194,191 | Denosumab<br>N = 4,276 | Oral<br>bisphosphonate<br>N = 75,293 |
| **Demographics** |  |  |  |  |
| **Age (years)** |  |  |  |  |
| Mean (SD) | 78 (10) | 74 (11) | 74 (10) | 71 (10) |
| **UK region** |  |  |  |  |
| East Midlands | 62 (0.9%) | 3,471 (1.8%) |  |  |
| East of England | 174 (2.7%) | 9,038 (4.7%) |  |  |
| London | 222 (3.4%) | 20,163 (10.4%) |  |  |
| North East | 344 (5.3%) | 6,985 (3.6%) |  |  |
| North West | 748 (11.5%) | 28,092 (14.5%) |  |  |
| Northern Ireland | 168 (2.6%) | 4,637 (2.4%) |  |  |
| Scotland | 528 (8.1%) | 17,474 (9.0%) |  |  |
| South Central | 1,296 (19.9%) | 18,999 (9.8%) |  |  |
| South East Coast | 683 (10.5%) | 16,909 (8.7%) |  |  |
| South West | 1,196 (18.3%) | 24,292 (12.5%) |  |  |
| Wales | 413 (6.3%) | 13,374 (6.9%) |  |  |
| West Midlands | 582 (8.9%) | 25,524 (13.1%) |  |  |
| Yorkshire And The Humber | 112 (1.7%) | 5,204 (2.7%) |  |  |
| Missing | 0 (0.0%) | 29 (0.0%) |  |  |
| **Denmark region** |  |  |  |  |
| Capital Region of Denmark |  |  | 1,185 (27.7%) | 19,118 (25.4%) |
| Central Denmark Region |  |  | 1,292 (30.2%) | 18,515 (24.6%) |
| North Denmark Region |  |  | 555 (13.0%) | 9,747 (12.9%) |
| Region of Southern Denmark |  |  | 779 (18.2%) | 17,466 (23.2%) |
| Region Zealand |  |  | * | * |
| Missing |  |  | * | * |
| **Socioeconomic status<sup>^</sup>** |  |  |  |  |
| 1 (most deprived) | 1,639 (25.1%) | 37,791 (19.5%) |  |  |
| 2 | 1,181 (18.1%) | 34,141 (17.6%) |  |  |
| 3 | 1,079 (16.5%) | 31,364 (16.2%) |  |  |
| 4 | 864 (13.2%) | 27,764 (14.3%) |  |  |
| 5 (least deprived) | 583 (8.9%) | 23,588 (12.1%) |  |  |
| Missing | 1,182 (18.1%) | 39,543 (20.4%) |  |  |
| **Family wealth<sup>^</sup>** |  |  |  |  |
| Low |  |  | 1,584 (37.0%) | 25,370 (33.7%) |
| Medium |  |  | 1,273 (29.8%) | 25,431 (33.8%) |
| High |  |  | 1,419 (33.2%) | * |
| Missing |  |  | 0 | * |
| **Education level<sup>^</sup>** |  |  |  |  |
| Low |  |  | 1,957 (47.1%) | 34,525 (47.1%) |
| Medium |  |  | 2,049 (49.3%) | 36,436 (49.7%) |
| High |  |  | 149 (3.6%) | 2,348 (3.2%) |
| Missing |  |  | 121 | 1,984 |
| **Cohabiting status^** |  |  |  |  |
| Alone |  |  | 2,537 (59.3%) | 40,458 (53.7%) |
| Cohabiting |  |  | 1,678 (39.2%) | 33,659 (44.7%) |
| Other |  |  | 61 (1.4%) | 1,176 (1.6%) |
| **Marital status^** |  |  |  |  |
| Single | 57 (4.7%) | 1,535 (5.5%) |  |  |
| Married | 493 (40.4%) | 10,802 (38.9%) | 1,928 (48.3%) | 37,466 (53.1%) |
| Widowed | 373 (30.6%) | 8,986 (32.3%) | 1,378 (34.6%) | 21,642 (30.7%) |
| Divorced | 9 (0.7%) | 542 (2.0%) | 682 (17.1%) | 11,466 (16.2%) |
| Separated | 13 (1.1%) | 492 (1.8%) |  |  |
| Engaged | * | 73 (0.3%) |  |  |
| Co-habiting | 10 (0.8%) | 167 (0.6%) |  |  |
| Remarried | 0 (0.0%) | 25 (0.1%) |  |  |
| More than one category recorded on the same date | 264 (21.6%) | 5,165 (18.6%) |  |  |
| Missing | 5,308 | 166,404 | 288 | 4,719 |
| **Year of index date** |  |  |  |  |
| 2011 | 108 (1.7%) | 29,313 (15.1%) | 767 (17.9%) | 13,197 (17.5%) |
| 2012 | 341 (5.2%) | 27,869 (14.4%) | 751 (17.6%) | 11,605 (15.4%) |
| 2013 | 574 (8.8%) | 27,606 (14.2%) | 568 (13.3%) | 10,814 (14.4%) |
| 2014 | 843 (12.9%) | 25,170 (13.0%) | 595 (13.9%) | 10,277 (13.6%) |
| 2015 | 1,063 (16.3%) | 22,423 (11.5%) | 627 (14.7%) | 9,848 (13.1%) |
| 2016 | 1,073 (16.4%) | 21,156 (10.9%) | 521 (12.2%) | 9,799 (13.0%) |
| 2017 | 1,198 (18.4%) | 20,183 (10.4%) | 447 (10.5%) | 9,753 (13.0%) |
| 2018 | 1,328 (20.3%) | 20,471 (10.5%) |  |  |
| **General health** |  |  |  |  |
| Anorexia | 51 (0.8%) | 837 (0.4%) | 8 (0.2%) | 35 (0.0%) |
| Antiepileptic | 802 (12.3%) | 18,333 (9.4%) | 353 (8.3%) | 4,651 (6.2%) |
| Antihypertensive | 3,737 (57.2%) | 107,755 (55.5%) | 2,435 (56.9%) | 40,179 (53.4%) |
| Antiparathyroid | 11 (0.2%) | 126 (0.1%) | 275 (6.4%) | 919 (1.2%) |
| Antipsychotic | 523 (8.0%) | 14,011 (7.2%) | 166 (3.9%) | 2,635 (3.5%) |
| Antithyroid | 30 (0.5%) | 1,144 (0.6%) | 93 (2.2%) | 1,515 (2.0%) |
| Anxiolytic | 551 (8.4%) | 16,566 (8.5%) | 532 (12.4%) | 7,107 (9.4%) |
| Ankylosing spondylitis | 34 (0.5%) | 943 (0.5%) | 7 (0.2%) | 69 (0.1%) |
| Asthma | 922 (14.1%) | 25,711 (13.2%) | 113 (2.6%) | 1,858 (2.5%) |
| Atopy | 158 (2.4%) | 5,523 (2.8%) | * | * |
| Atrial fibrillation | 524 (8.0%) | 12,637 (6.5%) | 322 (7.5%) | 4,338 (5.8%) |
| Cardiovascular disease | 693 (10.6%) | 15,899 (8.2%) | 436 (10.2%) | 5,901 (7.8%) |
| **Charlson score** |  |  |  |  |
| 0 | 3,553 (54.4%) | 107,715 (55.5%) | 2,729 (63.8%) | 52,607 (69.9%) |
| 1 | 1,668 (25.6%) | 51,036 (26.3%) | 911 (21.3%) | 15,622 (20.7%) |
| 2 | 736 (11.3%) | 19,983 (10.3%) | 333 (7.8%) | 4,352 (5.8%) |
| 3 | 368 (5.6%) | 10,399 (5.4%) | 143 (3.3%) | 1,466 (1.9%) |
| 4 | 127 (1.9%) | 3,394 (1.7%) | 91 (2.1%) | 723 (1.0%) |
| 5 | 52 (0.8%) | 1,127 (0.6%) | 47 (1.1%) | 318 (0.4%) |
| 6 | 17 (0.3%) | 400 (0.2%) | 11 (0.3%) | 122 (0.2%) |
| 7 | * | 110 (0.1%) | 9 (0.2%) | 57 (0.1%) |
| 8 | * | 21 (0.0%) | * | * |
| 9 | 0 (0.0%) | 6 (0.0%) | * | * |
| Chronic kidney disease | 970 (14.9%) | 24,904 (12.8%) | 127 (3.0%) | 532 (0.7%) |
| Chronic obstructive pulmonary disease | 766 (11.7%) | 21,694 (11.2%) | 493 (11.5%) | 6,465 (8.6%) |
| Colorectal polyps | 69 (1.1%) | 2,115 (1.1%) | * | * |
| Corticosteroid | 1,381 (21.2%) | 56,996 (29.4%) | 591 (13.8%) | 15,292 (20.3%) |
| **Crohn's disease** | 51 (0.8%) | 859 (0.4%) | 31 (0.7%) | 355 (0.5%) |
| **Dementia** | 457 (7.0%) | 8,936 (4.6%) | 111 (2.6%) | 1,206 (1.6%) |
| **Diabetes (any)** | 560 (8.6%) | 20,975 (10.8%) | 221 (5.2%) | 3,427 (4.6%) |
| **Direct factor Xa inhibitor** | 295 (4.5%) | 3,333 (1.7%) | 75 (1.8%) | 1,000 (1.3%) |
| **Direct thrombin inhibitor** | 28 (0.4%) | 367 (0.2%) | 60 (1.4%) | 628 (0.8%) |
| **Eczema** | 50 (0.8%) | 1,887 (1.0%) | 62 (1.4%) | 636 (0.8%) |
| **Heparin** | 108 (1.7%) | 2,295 (1.2%) | 13 (0.3%) | 207 (0.3%) |
| **Hormone replacement therapy** | 63 (1.0%) | 3,901 (2.0%) | 885 (20.7%) | 14,218 (18.9%) |
| **Hyperparathyroidism** | 98 (1.5%) | 2,068 (1.1%) | 114 (2.7%) | 1,534 (2.0%) |
| **Hyperlipidaemia** | 589 (9.0%) | 19,077 (9.8%) | 226 (5.3%) | 3,668 (4.9%) |
| **Hypocalcaemia** | 27 (0.4%) | 296 (0.2%) | 28 (0.7%) | 240 (0.3%) |
| **Hypoparathyroidism** | 0 (0.0%) | 23 (0.0%) | 147 (3.4%) | 1,597 (2.1%) |
| **Immunosuppressant** | 415 (6.4%) | 8,102 (4.2%) | 92 (2.2%) | 2,707 (3.6%) |
| **Inflammatory bowel disease/ulcerative colitis** | 42 (0.6%) | 1,215 (0.6%) | 43 (1.0%) | 585 (0.8%) |
| **Intravenous bisphosphonate** | 18 (0.3%) | 18 (0.0%) | 0 (0.0%) | 0 (0.0%) |
| **Ischaemic heart disease** | 274 (4.2%) | 6,617 (3.4%) | 390 (9.1%) | 4,783 (6.4%) |
| **Kyphosis** | 76 (1.2%) | 735 (0.4%) | 5 (0.1%) | 52 (0.1%) |
| **Liver cirrhosis** | 41 (0.6%) | 874 (0.5%) | 28 (0.7%) | 226 (0.3%) |
| **Lupus** | 27 (0.4%) | 595 (0.3%) | 16 (0.4%) | 154 (0.2%) |
| **Malnutrition** | 9 (0.1%) | 135 (0.1%) | 14 (0.3%) | 96 (0.1%) |
| **Menopausal symptoms** | 75 (1.1%) | 4,893 (2.5%) | 54 (1.3%) | 1,189 (1.6%) |
| **Multiple sclerosis** | 37 (0.6%) | 691 (0.4%) | 31 (0.7%) | 395 (0.5%) |
| **Non-steroidal anti-inflammatory drug** | 794 (12.2%) | 40,577 (20.9%) | 996 (23.3%) | 23,111 (30.7%) |
| **Number of different drugs** |  |  |  |  |
| Median (IQR) | 13 (8 - 19) | 10 (6 - 16) | 7 (4 - 11) | 6 (3 - 10) |
| **Number of GP/outpatient visits** |  |  |  |  |
| Median (IQR) | 19 (11 - 30) | 16 (9 - 27) | 2 (1 - 5) | 1 (0 - 3) |
| **Number of hospital admissions** |  |  |  |  |
| Median (IQR) | 0 (0 - 1) | 0 (0 - 1) | 0 (0 - 1) | 0 (0 - 1) |
| **Opioid** | 2,758 (42.2%) | 82,205 (42.3%) | 1,572 (36.8%) | 24,924 (33.1%) |
| **Osteoarthritis** | 1,331 (20.4%) | 36,155 (18.6%) | 524 (12.3%) | 8,162 (10.8%) |
| **Overweight/obesity** | 189 (2.9%) | 8,383 (4.3%) | 64 (1.5%) | 1,317 (1.7%) |
| **Parkinson's disease** | 121 (1.9%) | 1,743 (0.9%) | 45 (1.1%) | 402 (0.5%) |
| **Platelet aggregation inhibitor** | 1,597 (24.5%) | 44,523 (22.9%) | 1,191 (27.9%) | 17,864 (23.7%) |
| **Proton pump inhibitor** | 3,764 (57.7%) | 88,704 (45.7%) | 1,702 (39.8%) | 20,943 (27.8%) |
| **Psoriasis** | 114 (1.7%) | 4,215 (2.2%) | 14 (0.3%) | 247 (0.3%) |
| **Peripheral vascular disease** | 84 (1.3%) | 1,621 (0.8%) | 184 (4.3%) | 2,468 (3.3%) |
| **Rheumatoid arthritis** | 615 (9.4%) | 9,806 (5.0%) | 112 (2.6%) | 1,886 (2.5%) |
| **Sedatives** | 562 (8.6%) | 16,336 (8.4%) | 765 (17.9%) | 11,001 (14.6%) |
| **Selective estrogen receptor modulator** | 59 (0.9%) | 600 (0.3%) | 103 (2.4%) | 407 (0.5%) |
| **Selective serotonin reuptake inhibitor** | 923 (14.1%) | 28,024 (14.4%) | 547 (12.8%) | 7,982 (10.6%) |
| **Serious infection** | 3,522 (54.0%) | 97,530 (50.2%) | 1,100 (25.7%) | 14,015 (18.6%) |
| **Statin** | 2,291 (35.1%) | 68,769 (35.4%) | 1,186 (27.7%) | 21,696 (28.8%) |
| **Stroke** | 305 (4.7%) | 6,506 (3.4%) | 154 (3.6%) | 2,140 (2.8%) |
| **Strontium** | 355 (5.4%) | 1,947 (1.0%) | 203 (4.7%) | 338 (0.4%) |
| **Teriparatide** | 31 (0.5%) | 20 (0.0%) | 245 (5.7%) | 816 (1.1%) |
| **Thyroid disorder** | 528 (8.1%) | 14,801 (7.6%) | 0 (0.0%) | 0 (0.0%) |
| **Tricyclic antidepressant** | 980 (15.0%) | 26,996 (13.9%) | 174 (4.1%) | 2,122 (2.8%) |
| **Vitamin K inhibitor** | 397 (6.1%) | 10,810 (5.6%) | 238 (5.6%) | 3,400 (4.5%) |
| **Fractures and falls** |  |  |  |  |
| **Calcium/vitamin D** | 4,510 (69.1%) | 49,566 (25.5%) | 117 (2.7%) | 1,001 (1.3%) |
| **Cumulative oral bisphosphonate use (days)** |  |  |  |  |
| Mean (SD) | 241 (384) | 15 (102) | 193 (340) | 10 (86) |
| Median (IQR) | 0 (0 - 340) | 0 (0 - 0) | 0 (0 - 200) | 0 (0 - 0) |
| **Fall** | 1,576 (24.1%) | 33,722 (17.4%) |  |  |
| **Fracture** | 3,374 (51.7%) | 74,950 (38.6%) | 1,058 (24.7%) | 16,888 (22.4%) |
| **Number of fractures** |  |  |  |  |
| Mean (SD) | 1 (1) | 1 (1) | 0.34 (0.68) | 0.28 (0.58) |
| **Number of fracture types** |  |  |  |  |
| No fracture | 3,154 (48.3%) | 119,241 (61.4%) | 3,218 (75.3%) | 58,405 (77.6%) |
| 1 | 2,253 (34.5%) | 58,443 (30.1%) | 887 (20.7%) | 14,862 (19.7%) |
| 2 | 1,058 (16.2%) | 16,202 (8.3%) | 159 (3.7%) | 1,922 (2.6%) |
| 3 | 63 (1.0%) | 305 (0.2%) | 12 (0.3%) | 104 (0.1%) |
| **Number of days since last fracture** |  |  |  |  |
| Median (IQR) | 363 (162 - 799) | 88 (40 - 207) | 526 (210 - 1,037) | 227 (96 - 757) |
| **Vitamin D deficiency** | 522 (8.0%) | 7,532 (3.9%) | 56 (1.3%) | 765 (1.0%) |

CPRD: Clinical Practice Research Datalink; DNR: Danish National Registries; IQR: interquartile range; SD: standard deviation
*Results presented in number (%) unless indicated otherwise*
*\* Numbers masked when less than minimum cell count of 5.*
*^Socioeconomic status represented by the Index of Multiple Deprivation in the UK, and by family wealth, education level, cohabiting status, and marital status in Denmark*

**Table 2:** Number of imbalanced covariates in new user cohorts.

|  | CPRD | DNR |
| --- | --- | --- |
|  | n/N, (%) of imbalanced covariates | n/N, (%) of imbalanced covariates |
| Prior to PS estimation | 22/87, (25.3%) | 18/83, (21.7%) |
| PS matching | 3/87, (3.4%) | 4/83, (4.2%) |
| PS stratification | 3/87, (3.4%) | 9/83, (10.8%) |
| IPTW | 10/87, (11.5%) | 2/83 (2.4%) |

CPRD: Clinical Practice Research Datalink; DNR: Danish National Registries; IPTW: inverse probability of treatment weighting; PS: propensity score
N refers to the number of covariates in the PS model; n refers to the number of imbalanced covariates after PS methods

Eighty-seven covariates were used to estimate the PS. The median (interquartile range (IQR)) PS amongst denosumab and bisphosphonate users were 0.123 (0.048, 0.303) and 0.008 (0.004, 0.023), respectively. Figure S1 illustrates the PS distribution is right skewed in both treatment groups however, there were fewer bisphosphonate users with PS above 0.4.

##### PS matching

The PS matching algorithm selected 5,837 denosumab patients and 22,393 bisphosphonate patients thus excluding 691 (11%) denosumab users and 171,798 (88%) bisphosphonate users. After matching, the PS distribution was identical in the two treatment groups (Figure S1). Three (3.4%) covariates remained imbalanced: cumulative oral bisphosphonate use, vitamin D deficiency, and factor Xa inhibitor prescription (Figure S2).

In NCO analysis, 19 NCOs had at least 100 events. The effect of treatment on each NCO is shown in Figure S3. For Approach 1, four (21.1%) of the NCOs that were analysed had 95% CI excluding one (bowel incontinence, delirium, early fracture, and ingrown toenail). For approach 2, only one (5.3%) NCO, ingrown toenail (HR 2.18, 95% CI: 1.49, 3.21), had its 95% CI excluding one and its lower CI bound >1.30.

##### PS stratification

The PS distribution amongst denosumab and bisphosphonate users were similar in the first seven strata. In the 8^th^, 9^th^ and 10^th^ strata, the median and IQR was slightly larger for denosumab users than bisphosphonate users. Overall, within stratum there was good PS overlap between the treatment groups (Figure S4). On average, three covariates had ASMD >0.1: cumulative oral bisphosphonate use, recency of fracture, and strontium prescription (Figure S5).

There were 47 NCOs with at least 100 events and the effect of treatment on NCO is shown in Figure S6. Approach 1 had eight (17.0%) NCOs (atelectasis, bowel incontinence, delirium, early fracture, ingrown toenail, ankle sprain, strabismus, total hip arthroplasty due to osteoarthritis) with 95% CI excluding one. Approach 2 had three (6.4%) NCOs with 95% CI upper bound <0.75 (ankle sprain) or lower bound >1.30 (ingrown toenail and strabismus).

##### IPTW

The median (IQR) weights were 0.48 (0.11, 0.68) and 0.98 (0.97, 0.99) amongst the denosumab and bisphosphonate users, respectively. In the pseudo population, 10 covariates (11.5%) had ASMD >0.1: age, calcium/vitamin D prescription, cumulative oral bisphosphonate use, number of fractures, non-steroidal anti-inflammatory drug prescription, number of different drugs, proton pump inhibitor prescription, recency of fracture, region, and strontium prescription (Figure 2a).

**Figure 2a:**
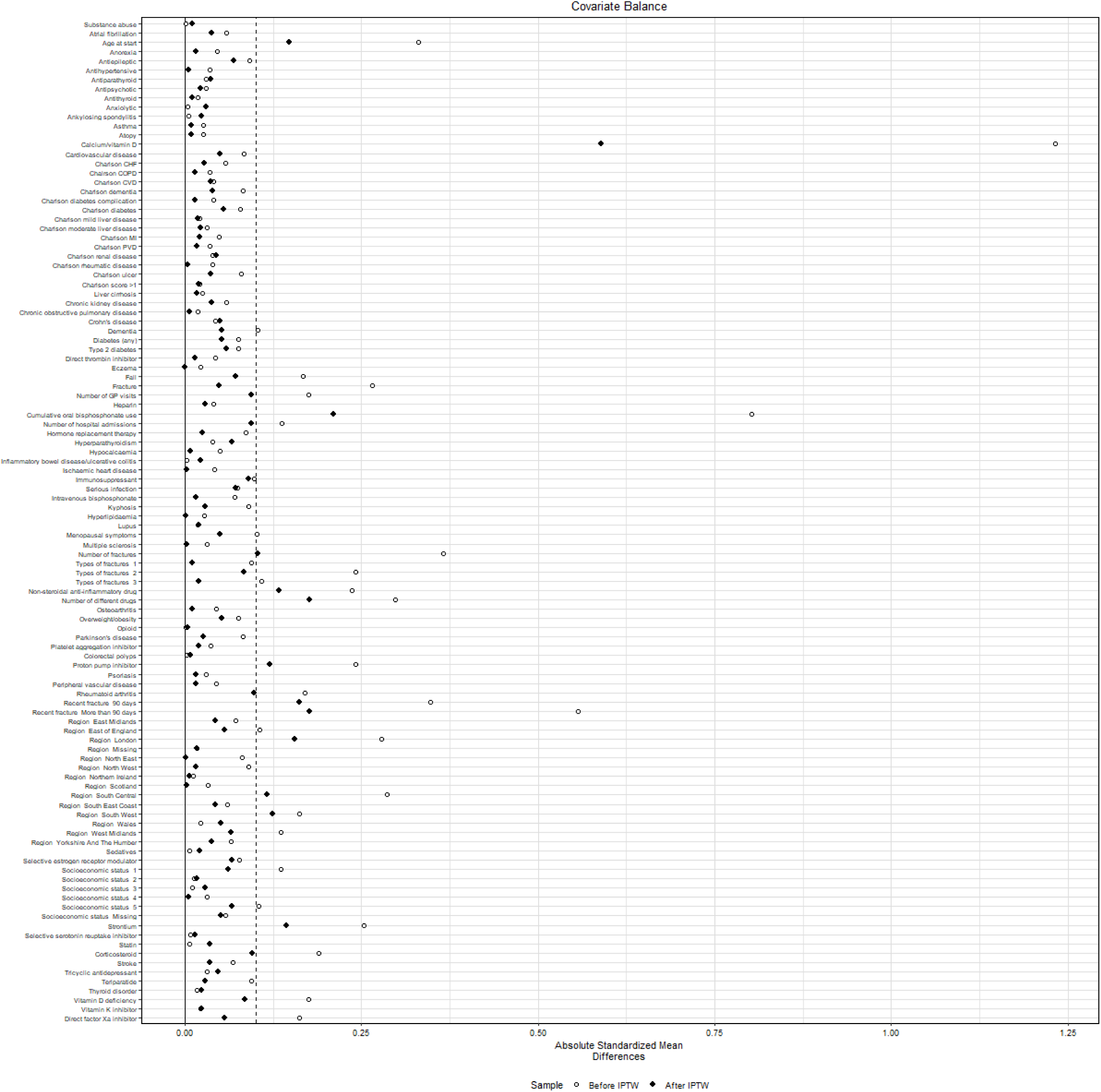
Covariate balance with propensity score weighting in new users (CPRD)

There were 47 NCOs with at least 100 events. In Approach 1, nine (19.1%) NCOs had its 95% CI excluding one (accident, anorectal disorder, delusional disorder, foreign body in ear, hypomagnesemia, ingrown nail, iron deficiency, nasal congestion, and schizophrenia). In Approach 2, two (4.3%) NCOs had its CI upper bound <0.75 (accident (0.03 (<0.01, 0.19)) and foreign body in ear (0.11 (0.02, 0.58)) (Figure 3a).

#### New switcher cohort

In CPRD, the new switcher study design analysed a smaller number of patients, 2,792 new denosumab switchers, and 14,668 new bisphosphonate switchers. In the pseudo-population, a smaller number of imbalanced covariates (2.4%, 2/82), as compared to the new user population, were observed which included previous fracture and cumulative oral bisphosphonate use (Figure S13). Thirteen NCOs had at least 100 events, of which none met the criteria for residual confounding using Approach 2 (Figure S14).

### DNR

#### New user cohort

DNR was a smaller database containing 79,569 eligible patients of which there were 4,276 denosumab users and 75,293 bisphosphonate users (Table 1). Most covariates were balanced between the two treatment groups; however, 18 (21.7%) covariates were imbalanced (Table 2), a smaller percentage compared to CPRD. Imbalanced patient characteristics were similar to CPRD, except higher prescription of antiparathyroid in denosumab users in DNR. (Table 1).

Eighty-three covariates were used to estimate the PS. The median (IQR) PS amongst denosumab and bisphosphonate users was 0.079 (0.044, 0.211) and 0.033 (0.023, 0.048). Figure S7 shows the PS distribution within treatment groups were right skewed however the distributions overlap each other. There were fewer bisphosphonate users with PS greater than 0.6.

##### PS matching

4,187 denosumab users were matched to 16,546 bisphosphonate users, excluding 89 (2%) denosumab and 58,747 (78%) bisphosphonate users. Four covariates (4.2%) remained imbalanced: number of outpatient visits, cumulative oral bisphosphonate use, number of prescriptions, and proton pump inhibitor use (Figure S8).

Twelve NCOs had at least 100 events with the effect of treatment on NCO shown in Figure S9. In Approach 1, only one (8.3%) NCO early fracture had its 95% CI excluding one. No NCO had met the criteria of Approach 2.

##### PS stratification

The PS distribution was similar between treatment groups in the first seven stratum. In the 8^th^, 9^th^ and 10^th^ strata, the median and IQR was slightly larger for denosumab users than bisphosphonate users. Overall, there is good overlap of PS distributions between the treatment groups (Figure S10). Nine covariates (10.8%) were imbalanced (Figure S11): age, renal disease, Charlson comorbidity score, cohabitation status, number of GP visits, cumulative bisphosphonate use, number of prescriptions, region, and proton pump inhibitor use.

Twenty-six NCOs had at least 100 events and the effect of treatment on NCO is shown in Figure S12. In Approach 1, one (3.8%) NCO early fracture had its 95% CI excluding one. In Approach 2, no NCO had met the criteria.

##### IPTW

The median (IQR) of weights was 0.68 (0.25, 1.23) and 0.98 (0.97, 0.99) in denosumab and bisphosphonate users respectively. In the pseudo-population, two (2.4%) covariates, age, and cumulative oral bisphosphonate use, were imbalanced (Figure 2b).

**Figure 2b:**
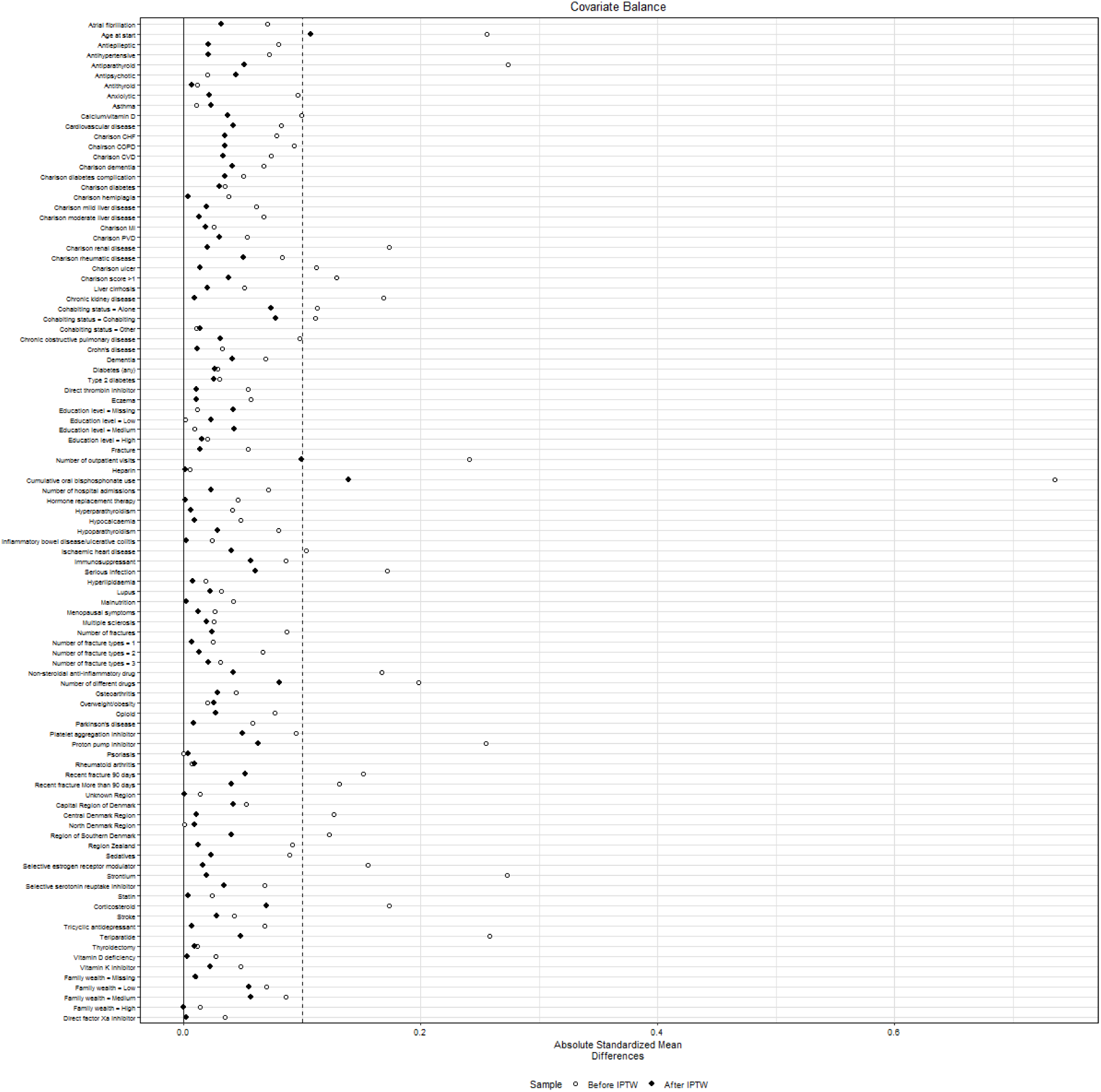
Covariate balance with propensity score weighting in new users (DNR)

Twenty-six NCOs had at least 100 events and the effect of treatment on NCO is shown in Figure 3b. In Approach 1, five (19.2%) NCOs had its 95% CIs excluding 1: anorectal disorder, eye injury, haematochezia, incomplete emptying of bladder, and total hip arthroplasty due to osteoarthritis. In Approach 2, no NCO had its 95% CI bound exceeding the indicated threshold of residual confounding.

**Figure 3a:**
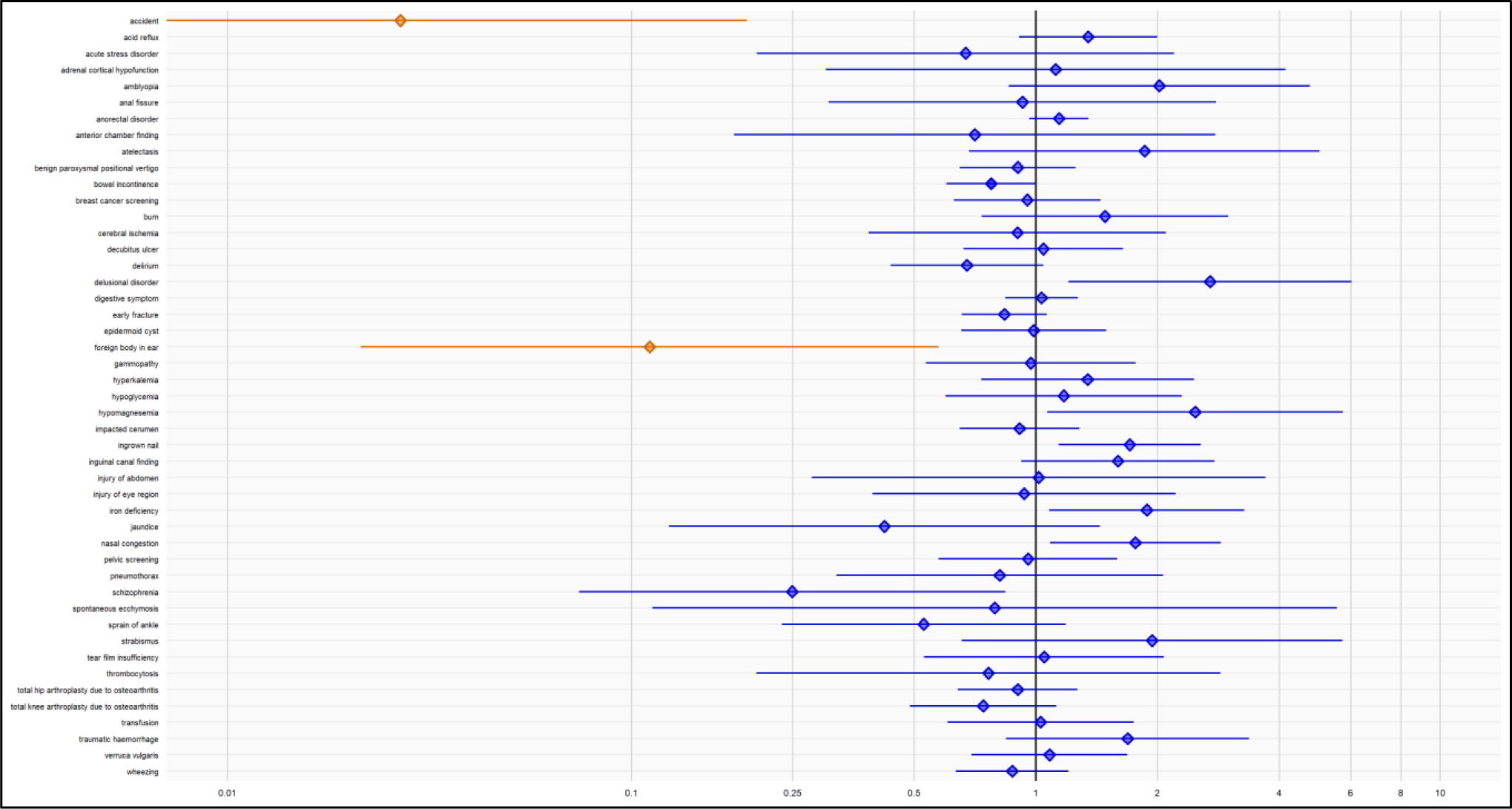
Negative control outcome estimates after PS weighting in new users (CPRD)

**Figure 3b:**
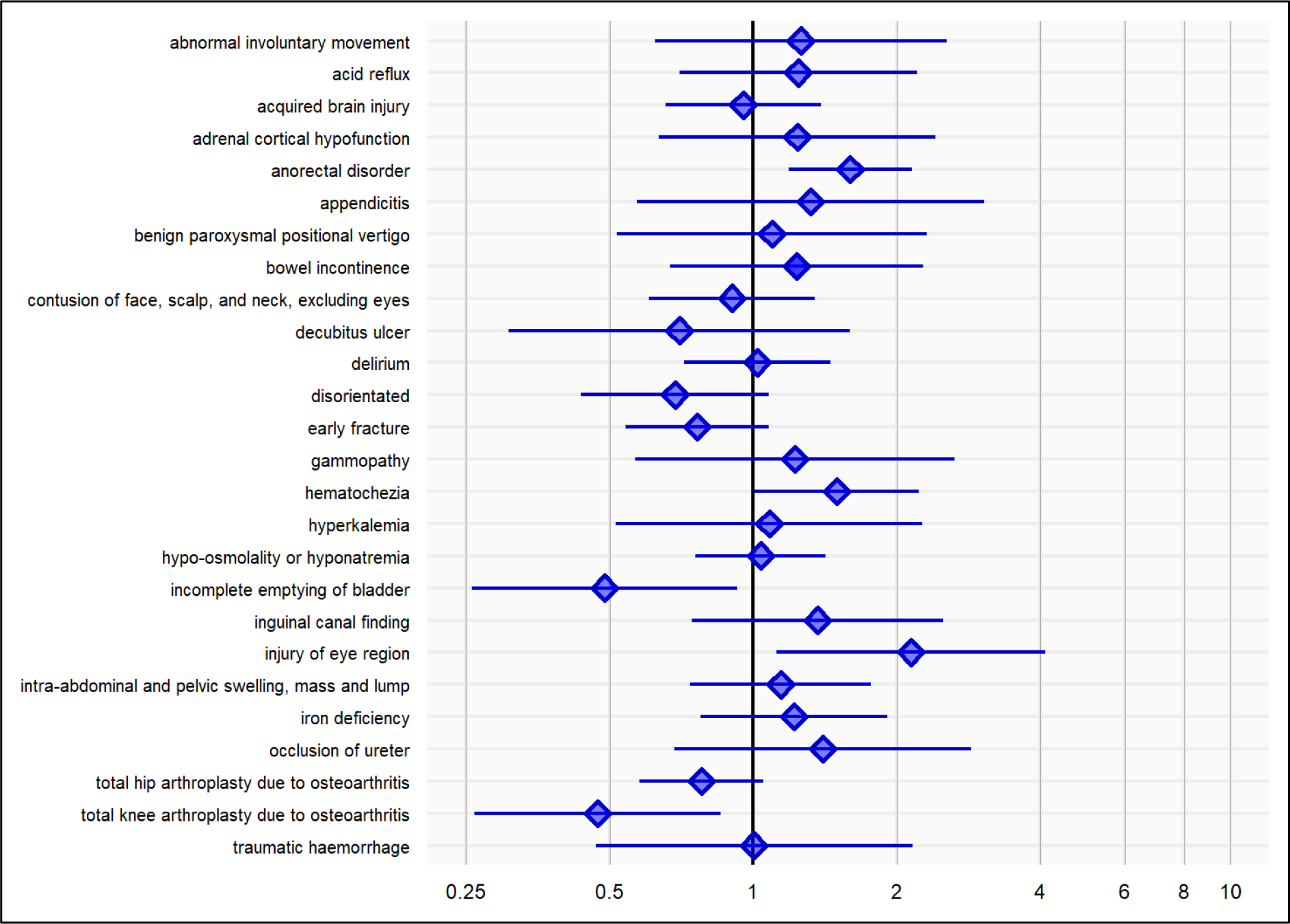
Negative control outcome estimates after PS weighting in new users (DNR)

#### New switcher cohort

In the new switcher cohorts, 3,726 new denosumab switchers and 2,525 new bisphosphonate switchers were eligible for the new switcher design. In the pseudo-population, a larger number of imbalanced covariates (5.1%, 4/78), as compared to the new user population, were observed which included previous fracture, number of fractures, number of fracture types, and recency of fracture (Figure S15). Comparability could not be assessed as there were no NCOs with at least 100 events.

#### Comparability assessment

After adjustment using PS methods, comparability of treatment groups based on meeting criteria for both Approach 1 and 2 was not achieved, with the exception of PS stratification in DNR (Table 3). Approach 1 was found to be more stringent than Approach 2 in both CPRD and DNR databases. Based on Approach 2, comparability was achieved for IPTW in CPRD, and all three PS methods in DNR. IPTW was considered preferable as it passed the criteria set in Approach 2 for both databases. The results for the subgroup analysis were similar (Tables S2 and S3). After IPTW, the treatment groups were comparable in the new switcher cohorts in CPRD.

**Table 3:** Comparability threshold using negative control outcome estimates in new user cohorts.

|  | CPRD |  |  | DNR |  |  |
| --- | --- | --- | --- | --- | --- | --- |
| Propensity score method | N | Approach 1<br>n (%) | Approach 2<br>n (%) | N | Approach 1<br>n (%) | Approach 2<br>n (%) |
| Matching | 19 | 4 (21.1) | 1 (5.3) | 12 | 1 (8.3) | 0 (0.0) |
| Stratification | 47 | 8 (17.0) | 3 (6.4) | 26 | 1 (3.8) | 0 (0.0) |
| Weighting | 47 | 9 (19.1) | 2 (4.3) | 26 | 5 (19.2%) | 0 (0.0) |

CPRD: Clinical Practice Research Datalink; DNR: Danish National Registries
N refers to number of negative control outcomes with at least 100 events during follow-up period. Results presented in number of negative control outcomes with residual bias [n (percentage)]

## Discussion

### Key results

This study had aimed to determine whether cohorts of patients selected for pharmacological treatment for osteoporosis with denosumab or oral bisphosphonates are comparable with respect to measured and unmeasured confounding.

In the UK and Denmark, denosumab is prescribed as second line treatment if an oral bisphosphonate is not suitable. As expected, baseline differences between the two treatment groups were observed with denosumab users being older, previously on preventative fracture treatment, and generally in worse health than bisphosphonate users. PS methods were used to ensure treatment groups were balanced for a wide range of covariates. Each PS method performed reasonably well in creating balanced groups, although the degree of performance varied by country. IPTW performed the least well in CPRD but performed the best in DNR, whereas stratification performed the best in CPRD but the worst in DNR; PS matching performed reasonably well in both databases, although it reduced sample size.

Evidence of residual confounding was based on satisfying both rules: more than 5% of NCOs had its 95% CI excluding the null value of one (Approach 1) or the CI was wholly outside the range of 0.75 to 1.30 (Approach 2). Based on both approaches, in CPRD, all PS methods showed evidence of residual confounding suggesting the new user treatment groups were not comparable. In DNR, PS stratification but not PS matching or IPTW showed treatment groups were comparable. Using Approach 1, large sample sizes may lead to narrow 95% CI and limit the possibility of crossing the null, even when effect sizes are clinically insignificant. Therefore, evidence of residual confounding based on Approach 2 alone may be considered for future analyses of comparative effectiveness.

IPTW was chosen to proceed with further as this satisfied the threshold in Approach 2 for both data sets. The new switcher analysis in DNR was not possible due to a small number of patients; in CPRD, the treatment groups were shown to be comparable.

### Comparison with previous studies

PS and NCO methods are popular approaches to account for confounding; however, there is no consensus on which threshold to use to determine whether treatment groups are comparable.

In our study, the degree to which PS methods ensured treatment groups were comparable varied by method and country, with other studies also observing similar findings. Choice of PS cannot be generalised to all databases as it is highly dependent on the research objective, achieving covariate balance, and the type and size of treatment effect estimate of interest (marginal, conditional, average treatment effect, or average treatment effect of the treated) (34–36).

Studies have approached the issue of residual confounding in different ways. McGrath (13) had assessed the comparability of newly initiating denosumab vs. oral bisphosphonates after using IPTW and 12 pre-specified NCOs using US claims data, with residual confounding being evident if a meaningful, non-null effect was observed. That study had found comparability was not achieved as associations were found for two NCOs thus comparative effectiveness estimation could not proceed. In another US study, Kim et al (29) evaluated three early fracture NCOs and four non-fracture NCOs; and assessed residual confounding using relative (risk ratio <0.85 or >1.15) and absolute (risk difference > 0.01) measures. In other populations, Levintow (14) had evaluated the comparability of lipid-lowering drugs using 10 pre-defined NCOs, assuming comparability was achieved if the risk difference was close to the null effect of zero (although a range was not specified) and the 95% CI contained zero for all NCOs. Other studies had instead focused on identifying a large set of NCOs in order to calibrate treatment effect estimates for residual confounding (15, 30, 37, 38) with Hripcsak (15) specifying that 95% of NCOs were expected to have the null effect contained within its 95% CI. Most commonly, studies choose one (or few) NCO(s) that assume to have the same confounding structure between treatment and primary outcome, and if a non-null effect is observed then residual confounding is present (11).

In contrast, and a strength of our study, we had used a combination of methods. Firstly, early fracture was selected a priori as an NCO assuming it shared the same set of confounders as treatment and primary outcome of fragility fracture. Secondly, an automated method identified over 100 potential NCOs that were unlikely to be associated with treatment; however, the assumption of sharing the same set of confounders is unlikely to be met; use of a large set of NCOs with differing confounding structures may mitigate that assumption. Thirdly, the optimal number of NCOs may be dependent on whether one wants to simply detect the presence of residual confounding, or to use NCOs to calibrate treatment effect estimates for residual confounding which would require at least 30 NCOs (39). The use of a large number of NCOs in our analysis allows for this possibility.

### Strengths and limitations

Our study was performed in population-based databases in the UK and Denmark. There was some similarities and differences in the achievement of comparability between databases.

Although new-user analyses are generally preferred for comparative studies, the predominant use of denosumab as second-line therapy in both the UK and Denmark may have contributed to difficulties in achieving comparability and to inclusion of patients not fully representative of clinical practice. We therefore conducted a new-switcher analysis to account for prior use of oral bisphosphonates in the denosumab group. Correspondingly, comparability appeared easier to achieve in the CPRD new switcher analysis than the new-user analysis.

We assessed whether residual confounding was evident based on two approaches with differing levels of strictness. Choice of threshold carries risks for decision-making on when to proceed with comparative assessments: a threshold that is too stringent risks excluding the possibility of conducting analyses on sufficiently comparable cohorts; a threshold that is not stringent enough may lead to comparisons on non-comparable cohorts.

Although our study had identified over 100 potential NCOs, under half were used in analysis as some NCOs were rare (less than 100 events). The decision to exclude rare outcomes was justified as they were unlikely to have adequate power to detect a non-null effect contributing misleading evidence of no residual confounding. However, imposing a minimum number of events had led to some analyses with a small number of assessable NCOs, resulting in only one NCO comparison needing to show a significant difference to exceed the 5% threshold of residual confounding.

Decisions made using IPTW may have affected the way standard errors and in turn CIs were calculated. Firstly, robust standard errors were used to ensure NCO estimates were not biased due to either a potential misclassification of PS estimation or the Cox model (40); this approach is known to over-estimate standard errors (41). Secondly, use of truncated weights would have led to more stable weights thus leading to smaller standard errors. Lastly, adjusting for imbalanced covariates may have led to larger standard errors (34). It is unclear what the overall impact was on standard errors, whether they were over- or under-estimated and the impact this would have had when using CIs to assess whether residual confounding existed.

Despite these limitations, a key strength is that all three PS methods were considered thus offering the flexibility that any one method could be used for further comparative effectiveness analysis.

### Conclusions

Confounding by indication will always be observed in routinely collected medical record data and PS and NCO methods are important tools to account for such confounding. We have shown that assessment of comparability varies depending on the method of PS adjustment and definitions of residual bias. The extent to which residual confounding can be identified is unknown, and studies should consider more than one PS method to test robustness and identify the largest number of NCOs to give the greatest flexibility in detecting residual confounding. Further research is required to determine the optimal threshold to identify residual confounding.

### Ethical approval

Access to CPRD data was approved (Protocol #20_000206) according to CPRD’s research data governance framework.

The study was reported to the Danish Data Protection Agency through registration at Aarhus University (record: AU-2016-051-000001, serial number 880).

An informed consent or ethical approval is not required for studies based solely on existing registry data.

## Supporting information

NCO methods paper supplementary

## Acknowledgments

This study was funded by Amgen Inc. The funding source was involved in the study protocol and manuscript review.

This study is based in part on data from the Clinical Practice Research Datalink obtained under license from the UK Medicines and Healthcare products Regulatory Agency. The data is provided by patients and collected by the NHS as part of their care and support. The interpretation and conclusions contained in this study are those of the authors alone.

We thank Uffe Heide-Jørgensen from Aarhus University for his assistance in data management and statistical analysis of the Danish National Registries.

All authors approved the final version of the manuscript and agree to be accountable for the work.

## Data availability statement

Qualified researchers may request data from the deidentified and aggregated results of this study from the corresponding author. The data are not publicly available due to privacy or ethical restrictions.

