## Supplementary material for "Evaluating the comparability of osteoporosis treatments using propensity score and negative control outcome methods in UK and Denmark electronic health record databases": NCO methods paper supplementary

Table S1: List of negative control outcomes and covariates

| **Negative control outcomes** | **Covariates** |
| --- | --- |
|  | ***Demographics^b^*** |
| Any fragility fracture in the 3 months post index date^a^ | Age*^ |
| Abnormal involuntary movement | Ethnicity (CPRD) |
| Abnormal pupil | Socioeconomic status*^ |
| Accident | Body mass index (CPRD) |
| Acid reflux | Smoking status (CPRD) |
| Acquired brain injury | Alcohol intake (CPRD) |
| Acute stress disorder | Region* |
| Adrenal cortical hypofunction | ***Comorbidities^b^*** |
| Amblyopia | Ankylosing spondylitis* |
| Anal and rectal polyp | Anorexia* |
| Anal fissure | Asthma*^ |
| Anorectal disorder | Atopy* |
| Anterior chamber finding | Atrial fibrillation*^ |
| Appendicitis | Cardiovascular disease (myocardial infarction, stroke, angina, transient ischemic attack)*^ |
| Ascites | Charlson comorbidity*^ |
| Atelectasis | Chronic kidney disease*^ |
| Bacteremia | Chronic obstructive pulmonary disease*^ |
| Benign paroxysmal positional vertigo | Colorectal polyps* |
| Bladder dysfunction | Crohn’s disease*^ |
| Bowel incontinence | Cumulative oral bisphosphonate use*^ |
| Breast cancer screening | Dementia*^ |
| Burn | Diabetes*^ |
| Calcaneal spur | Eczema*^ |
| Calculus of lower urinary tract | Exposure to radiation |
| Cerebral ischemia | History of fracture (number, site, and recency)*^ |
| Colorectal screening | Hyperlipidaemia*^ |
| Complete bilateral paralysis | Hypertriglyceridemia |
| Congenital heart disease | Hypo/hyperparathyroidism*^ |
| Conjunctival hyperemia | Hypoalbuminemia |
| Contusion of face, scalp, and neck, excluding eyes | Hypocalcaemia*^ |
| Corneal endothelial finding | Inflammatory bowel disease/ ulcerative colitis*^ |
| Decubitus ulcer | Ischemic heart disease*^ |
| Delirium | Kyphosis* |
| Delusional disorder | Liver cirrhosis*^ |
| Dependent personality disorder | Lupus*^ |
| Digestive symptom | Malnutrition^ |
| Disorientated | Menopausal symptoms*^ |
| Dysarthria | Multiple sclerosis*^ |
| Encephalomyelopathy | Osteoarthritis*^ |
| Endometriosis | Overweight/obesity*^ |
| Epidermoid cyst | Parkinson’s disease*^ |
| Explosive personality disorder | Peripheral vascular disease* |
| Finding of bowel incontinence | Polycystic ovarian syndrome |
| Food poisoning | Previous fall* |
| Foreign body in ear | Psoriasis*^ |
| Gammopathy | Rheumatic fever |
| Gender identity disorder | Rheumatoid arthritis*^ |
| Hematochezia | Serious infections*^ |
| Hemochromatosis | Stroke*^ |
| Hemoglobinopathy | Substance abuse* |
| Hyperkalemia | Thyroid disorder* |
| Hypertrophic scar | Thyroidectomy^ |
| Hypertrophy of breast | Vitamin D deficiency*^ |
| Hypogammaglobulinemia | Ankylosing spondylitis* |
| Hypoglycemia | ***Healthcare use^c^*** |
| Hypomagnesemia | Number of GP visits* |
| Hypo-osmolality or hyponatremia | Number of hospitalisations/hospital outpatient visits*^ |
| Idiopathic peripheral autonomic neuropathy | ***Medications^c^*** |
| Impacted cerumen | Anticoagulants and antithrombotics*^ |
| Incomplete emptying of bladder | Antidepressants*^ |
| Incoordination | Antiepileptics*^ |
| Ingrown nail | Antihypertensive drugs*^ |
| Inguinal canal finding | Antiparathyroid drugs (includes calcitonin)*^ |
| Injury of abdomen | Antipsychotics*^ |
| Injury of eye region | Antithyroid drugs*^ |
| Intra-abdominal and pelvic swelling, mass and lump | Anxiolytics*^ |
| Iron deficiency | Calcium/vitamin D*^ |
| Jaundice | Immunosuppressants*^ |
| Ketoacidosis | Intravenous bisphosphonates* |
| Macular drusen | Non-steroidal anti-inflammatory drugs*^ |
| Mental retardation | Opioids*^ |
| Mohs surgery | Oral corticosteroids*^ |
| Nasal congestion | Proton pump inhibitors*^ |
| Noise effects on inner ear | Sedatives*^ |
| Obsessive compulsive personality disorder | Selective estrogen receptor modulators*^ |
| Occlusion of ureter | Statins*^ |
| Optic atrophy | Strontium ranelate*^ |
| Panhypopituitarism | Systemic hormone replacement therapy (including tibolone)*^ |
| Paranoid personality disorder | Teriparatide*^ |
| Pelvic screening | Number of prescriptions*^ |
| Personality disorder |  |
| Phobic disorder |  |
| Pneumothorax |  |
| Postviral fatigue syndrome |  |
| Presbyopia |  |
| Psychalgia |  |
| Ptotic breast |  |
| Regular astigmatism |  |
| Schizoaffective disorder |  |
| Schizophrenia |  |
| Somatization disorder |  |
| Spontaneous ecchymosis |  |
| Sprain of ankle |  |
| Strabismus |  |
| Tear film insufficiency |  |
| Thrombocytosis |  |
| Tic disorder |  |
| Total hip arthroplasty due to osteoarthritis |  |
| Total knee arthroplasty due to osteoarthritis |  |
| Transfusion |  |
| Traumatic haemorrhage |  |
| Urethral stricture |  |
| Urethritis |  |
| Urgent desire for stool |  |
| Verruca vulgaris |  |
| Vesicoureteric reflux |  |
| Visual test |  |
| Weakness of face muscles |  |
| Wheezing |  |
| Wound dehiscence |  |

^a^Any fracture included hip, vertebral, radius, ulna, wrist, humerus, pelvis, clavicle, and shoulder; ^b^measured in the five years prior to the index date; ^c^measured in the 12 months prior to the index date; *included in propensity score for CPRD; ^included in propensity score for NDR

Figure S1: Propensity score distribution before and after matching (CPRD new user cohort)

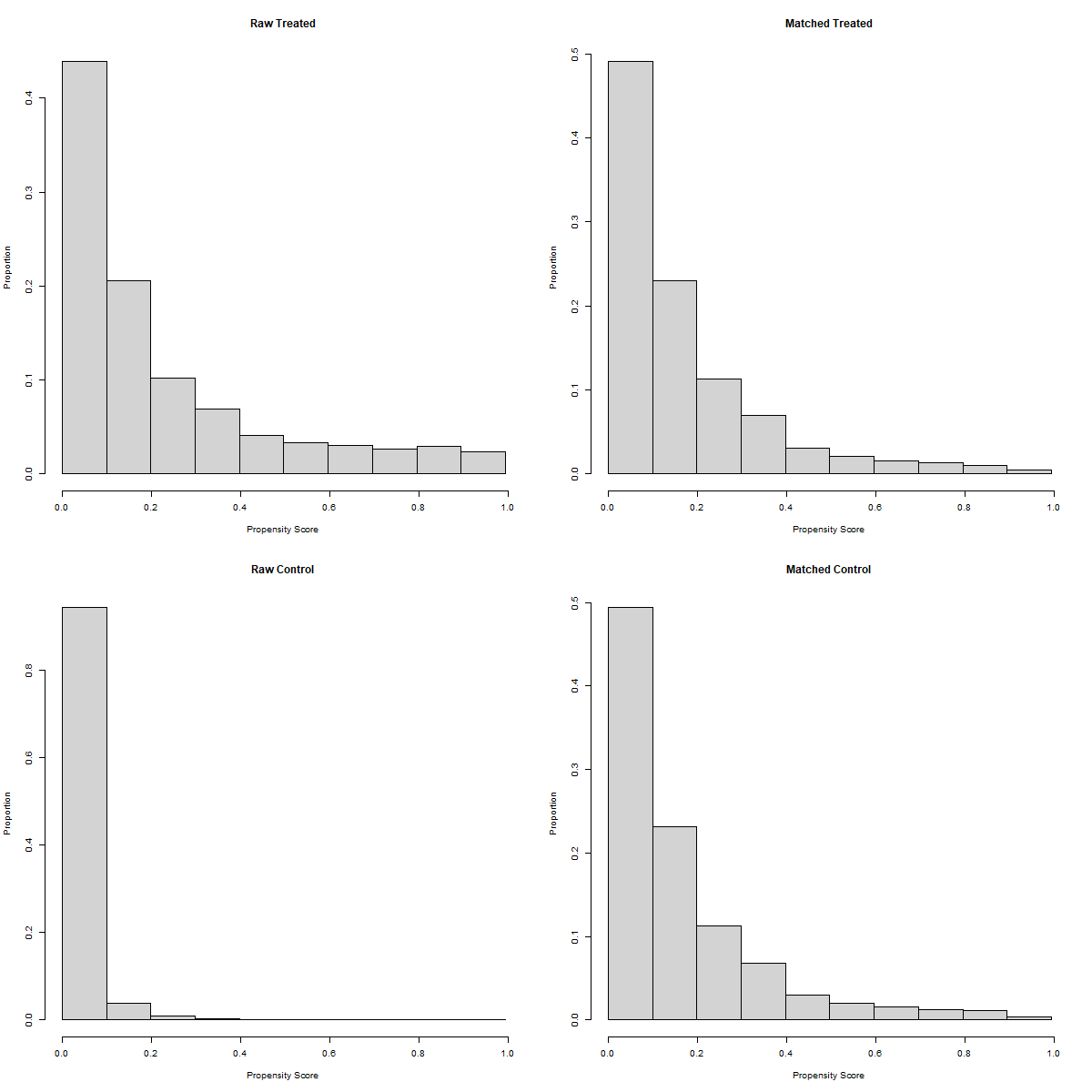

Figure S2: Covariate balance with propensity score matching (CPRD new user cohort)

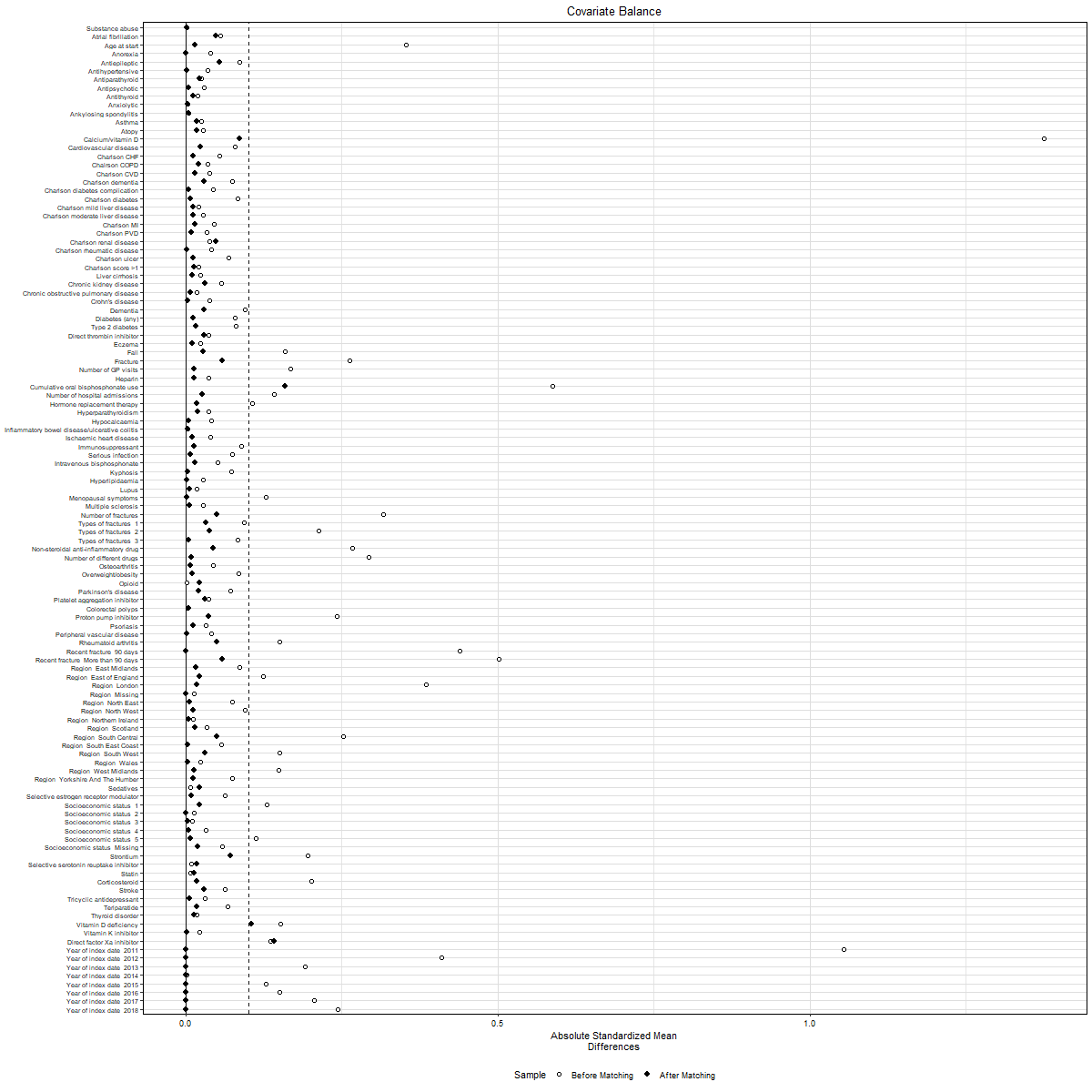

Figure S3: Negative control outcome estimates after PS matching (CPRD new user cohort)

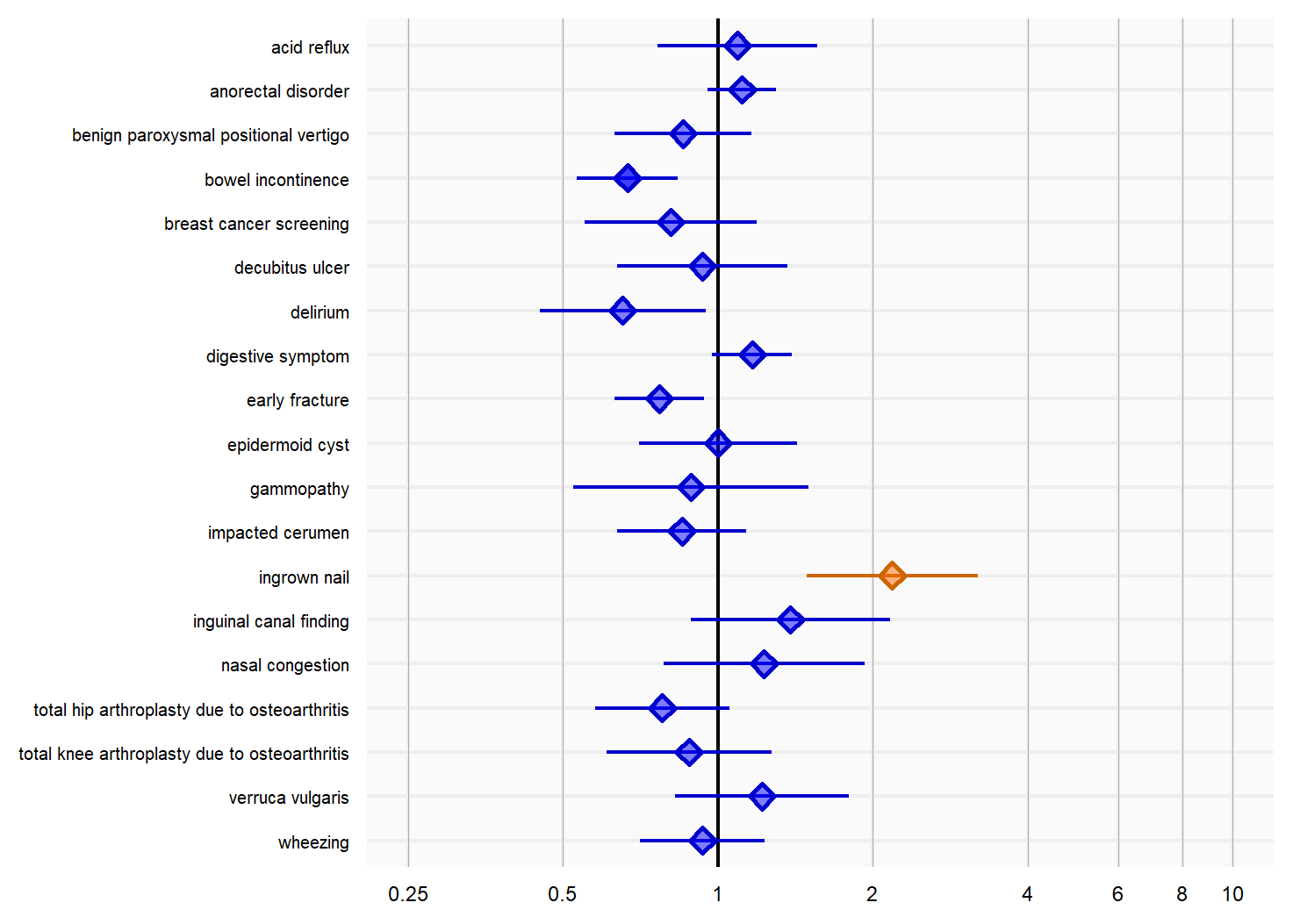

Figure S4: Propensity score distribution after stratification (CPRD new user cohort)

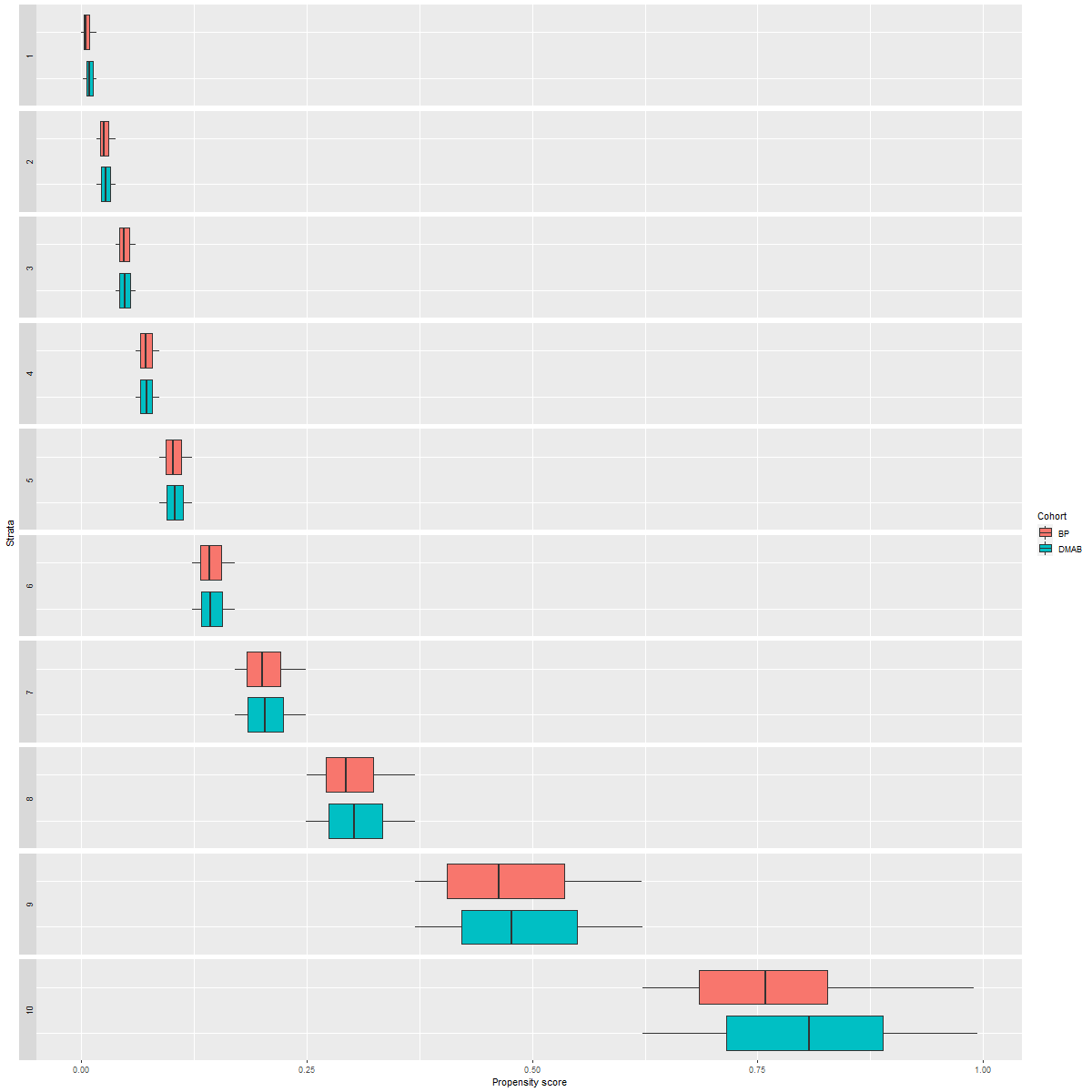

Figure S5: Covariate balance with propensity score stratification (CPRD new user cohort)

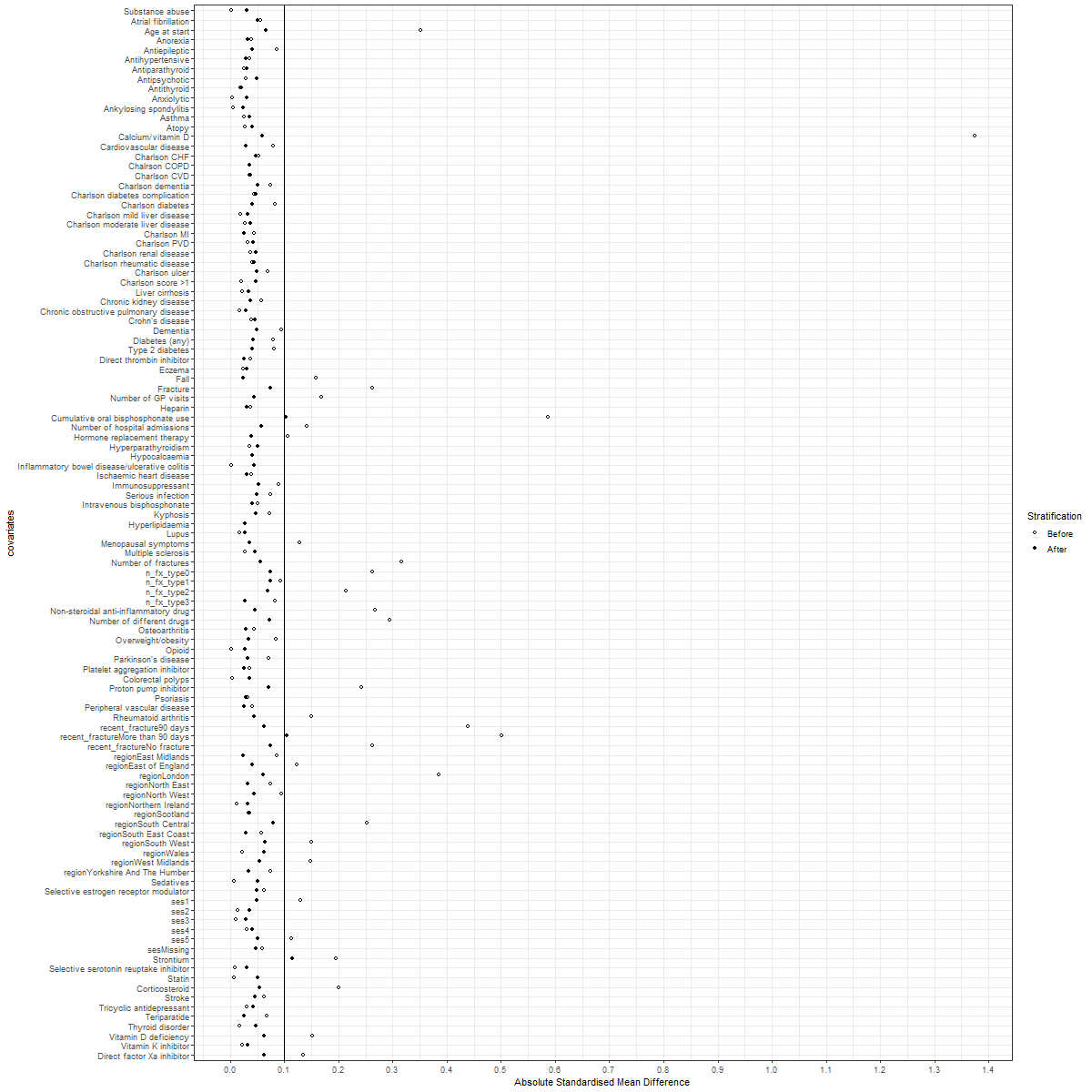

Figure S6: Negative control outcome estimates after PS stratification (CPRD new user cohort)

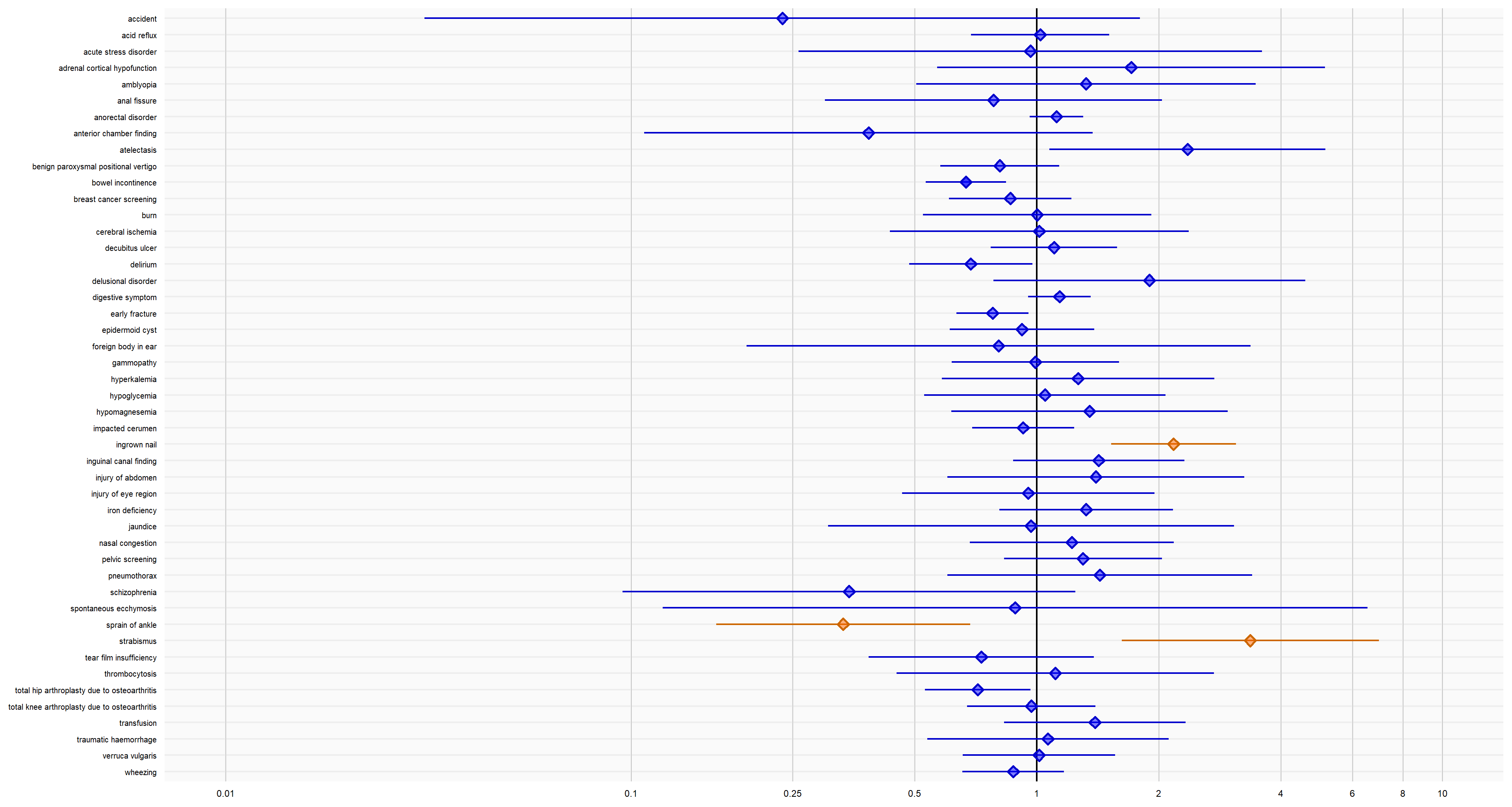

Figure S7: Propensity score distribution before and after matching (DNR new user cohort)

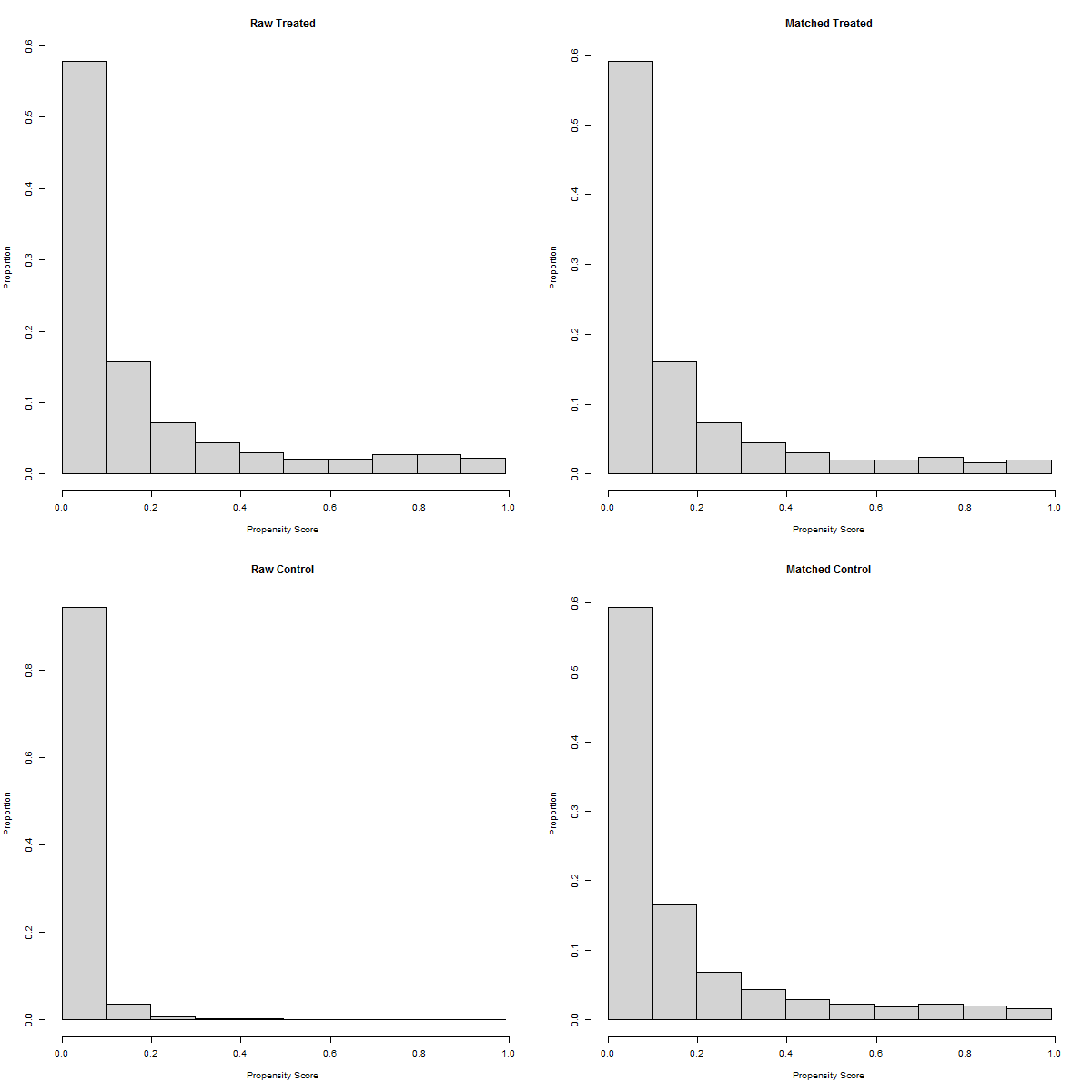

Figure S8: Covariate balance with propensity score matching (DNR new user cohort)

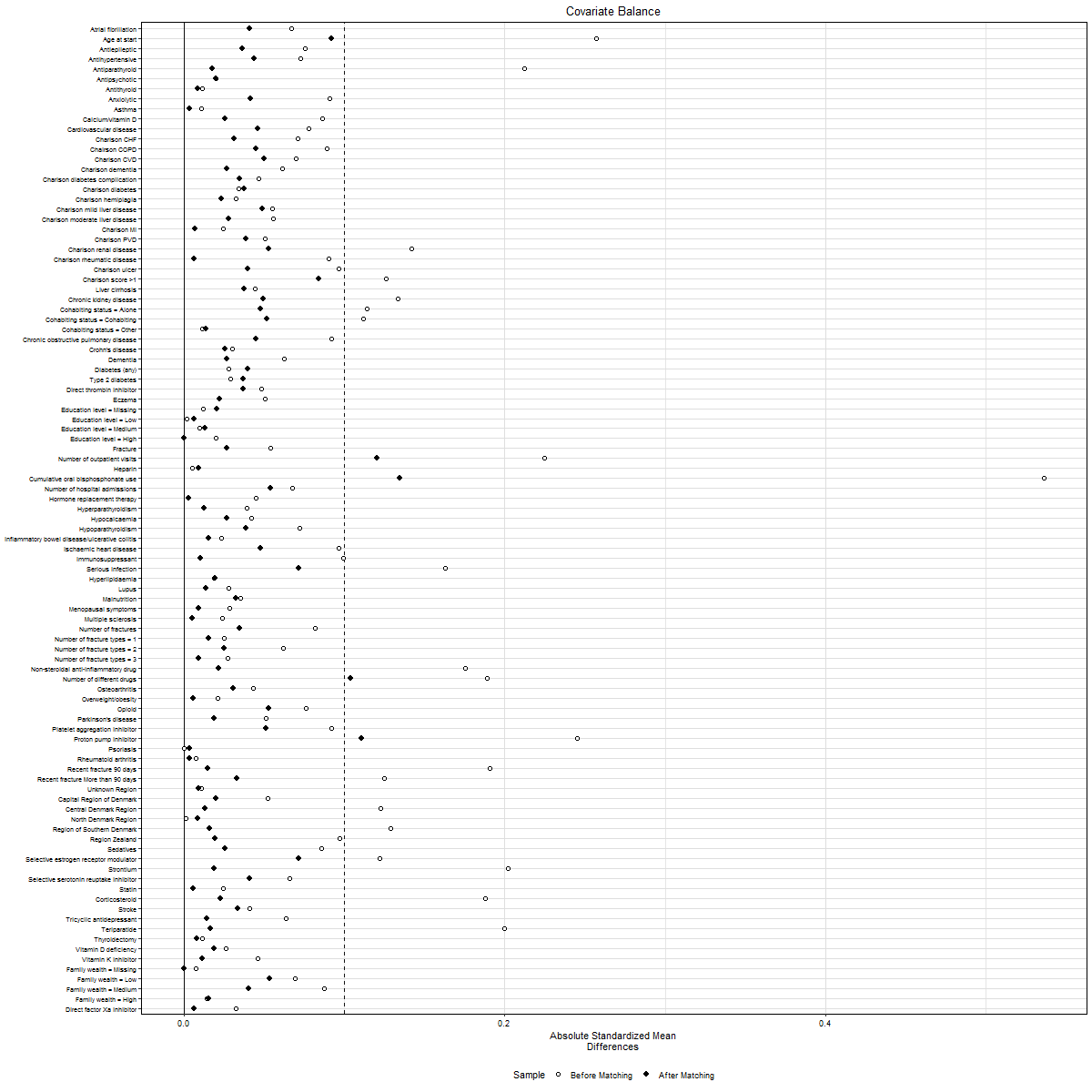

Figure S9: Negative control outcome estimates after PS matching (DNR new user cohort)

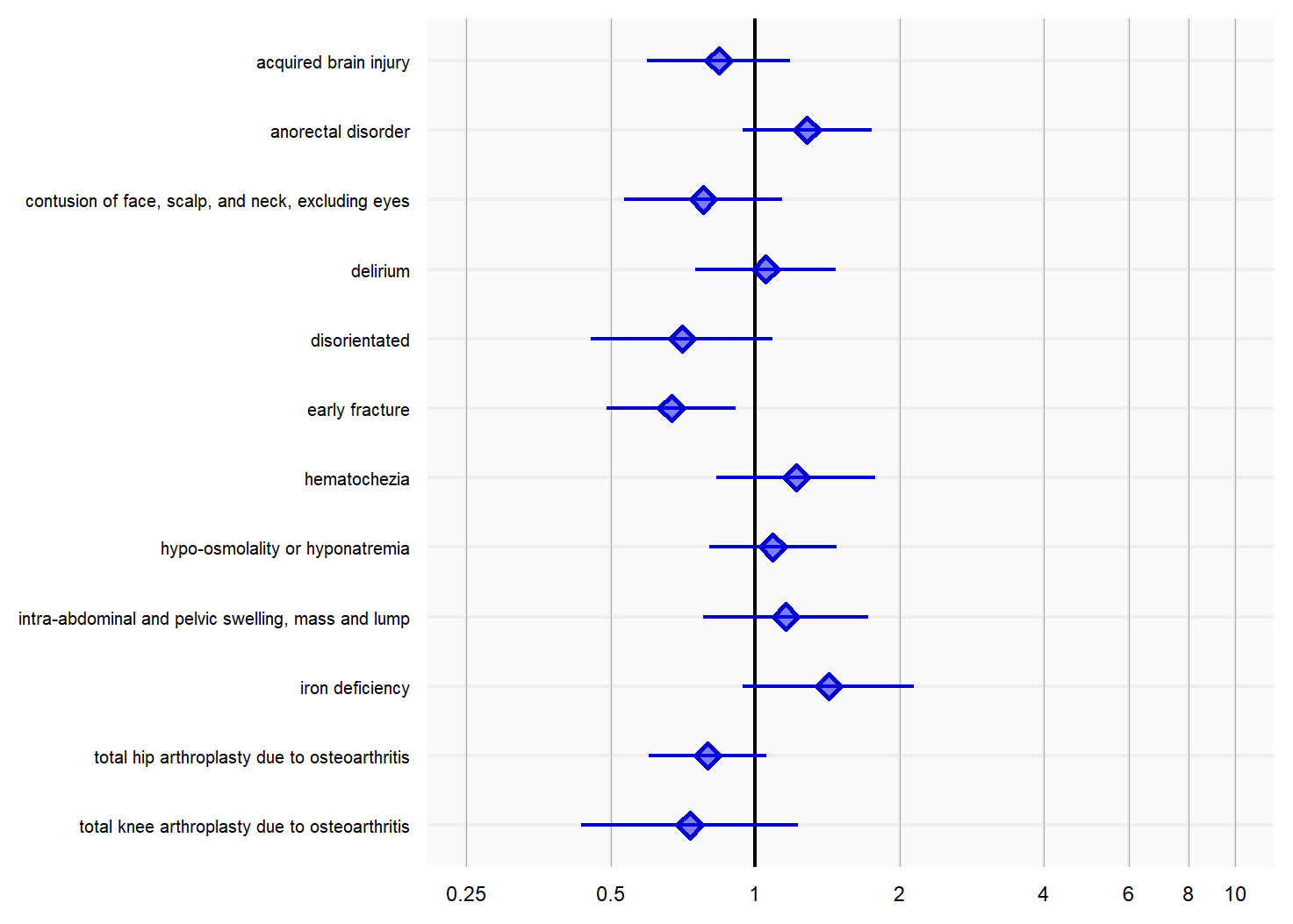

Figure S10: Propensity score distribution after stratification (DNR new user cohort)

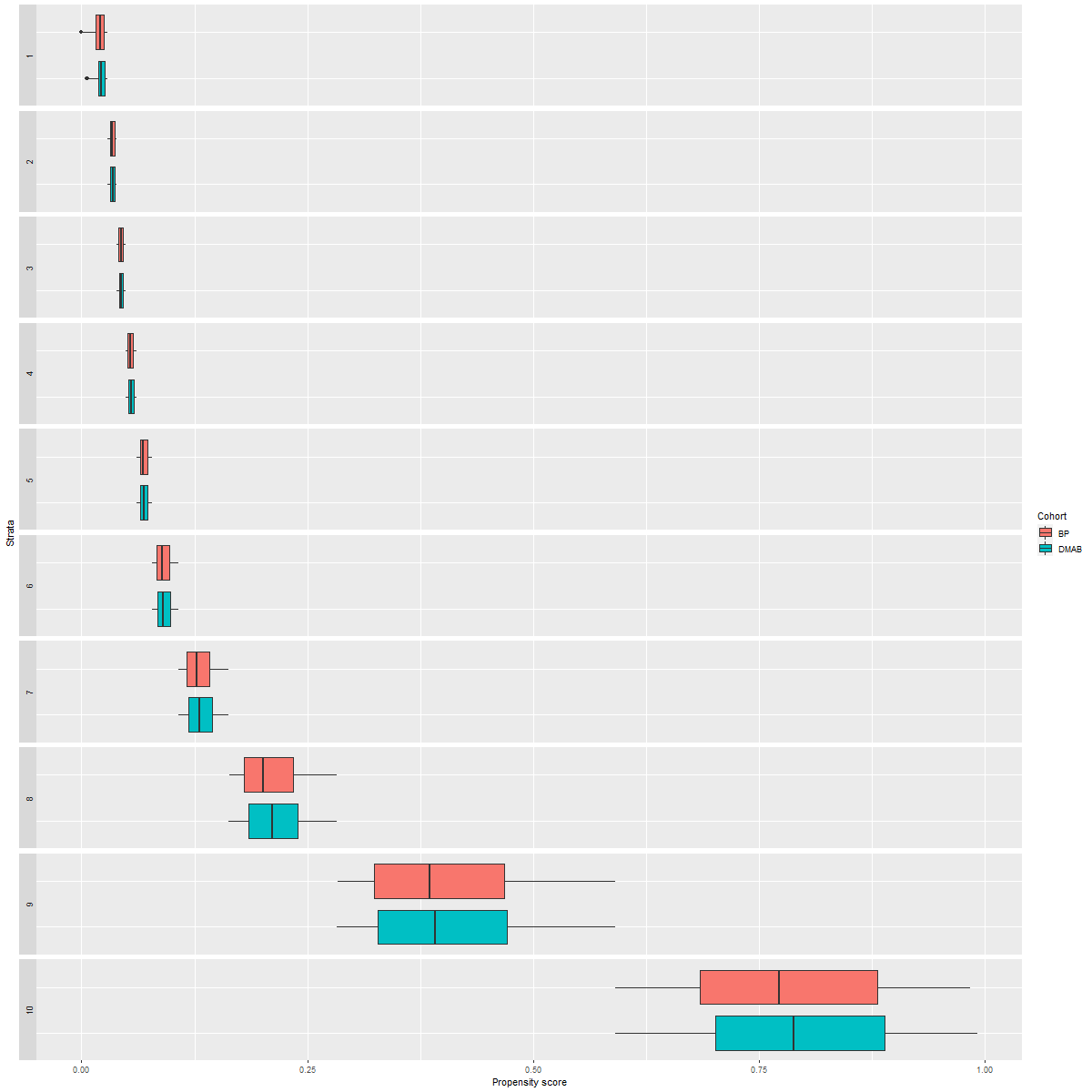

Figure S11: Covariate balance with propensity score stratification (DNR new user cohort)

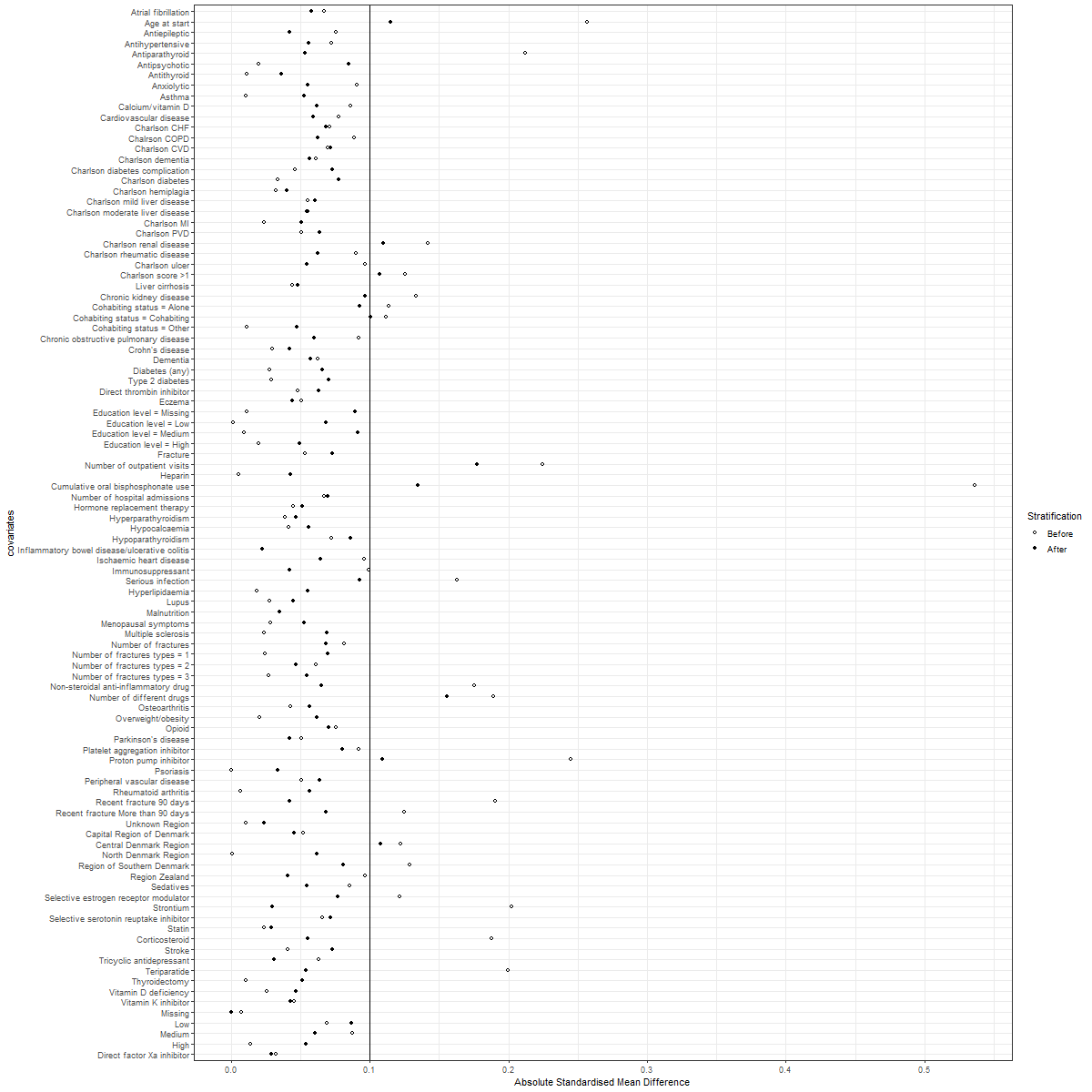

Figure S12: Negative control outcome estimates after PS stratification (DNR new user cohort)

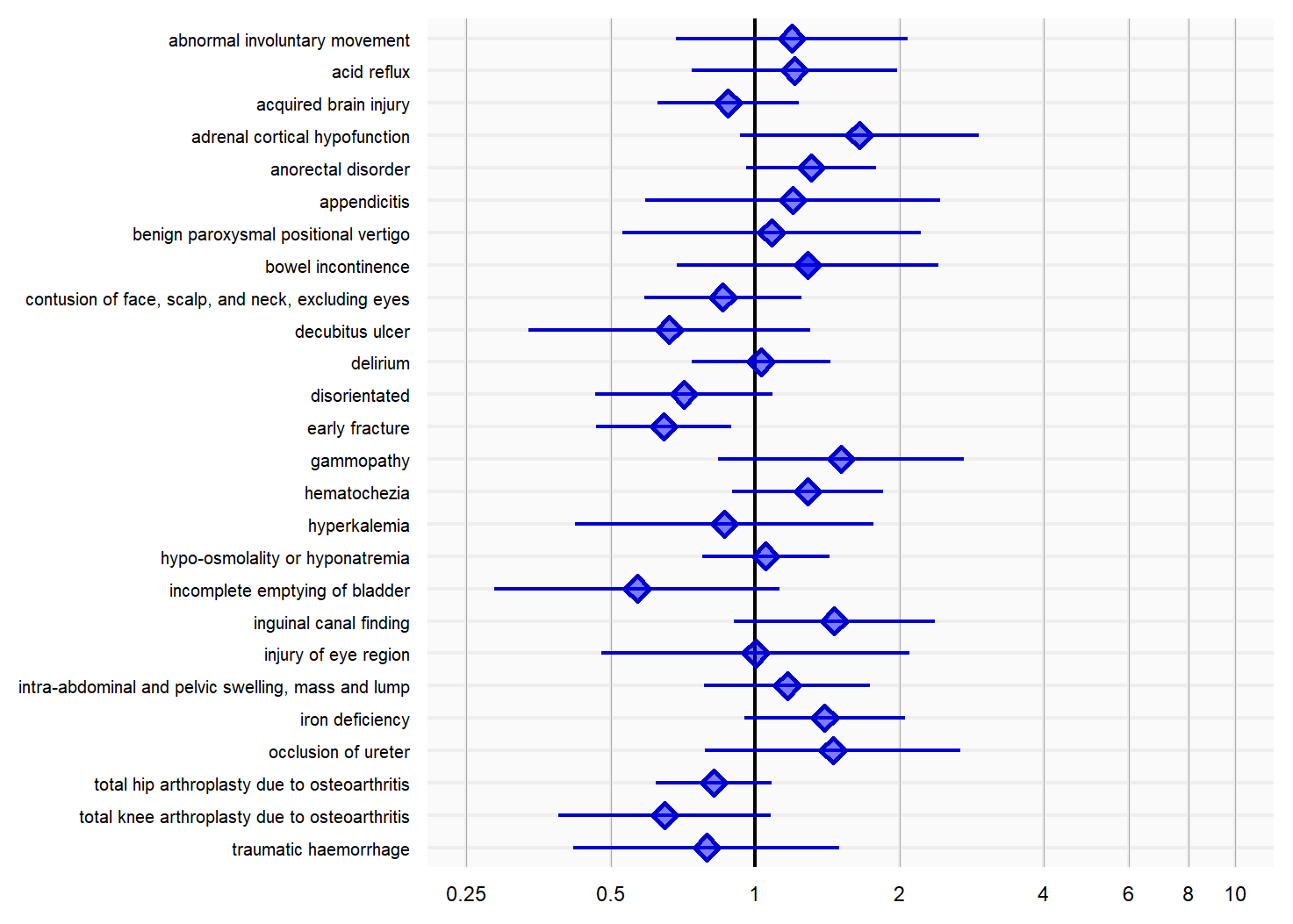

Figure S13: Covariate balance with PS weighting (CPRD new switcher cohort)

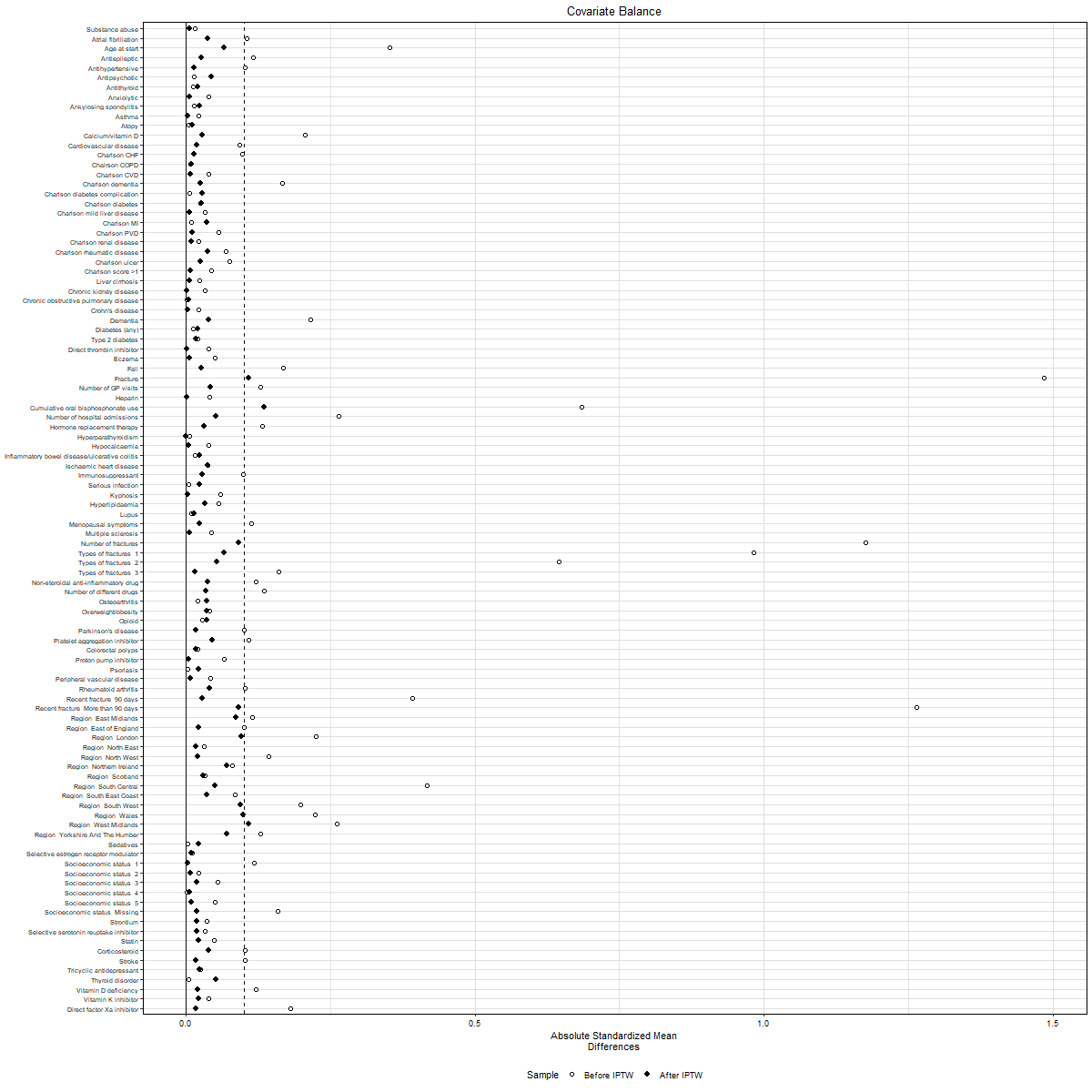

Figure S14: Negative control outcome estimates after PS weighting (CPRD new switcher cohort)

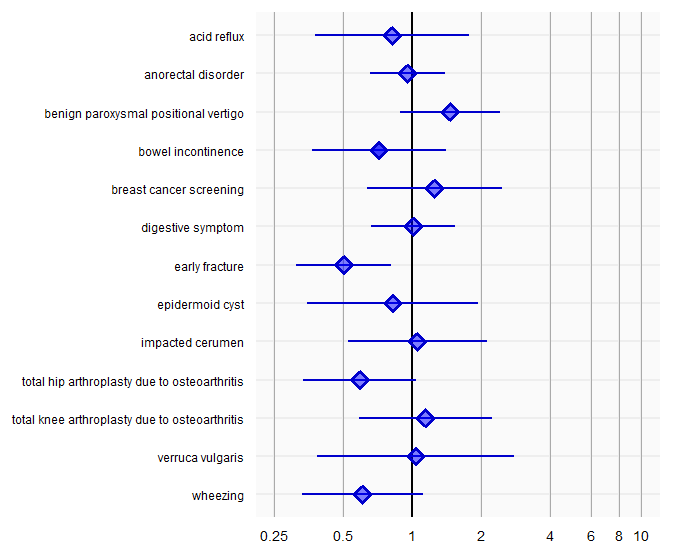

Figure S15: Covariate balance with PS weighting in (DNR new switcher cohort)

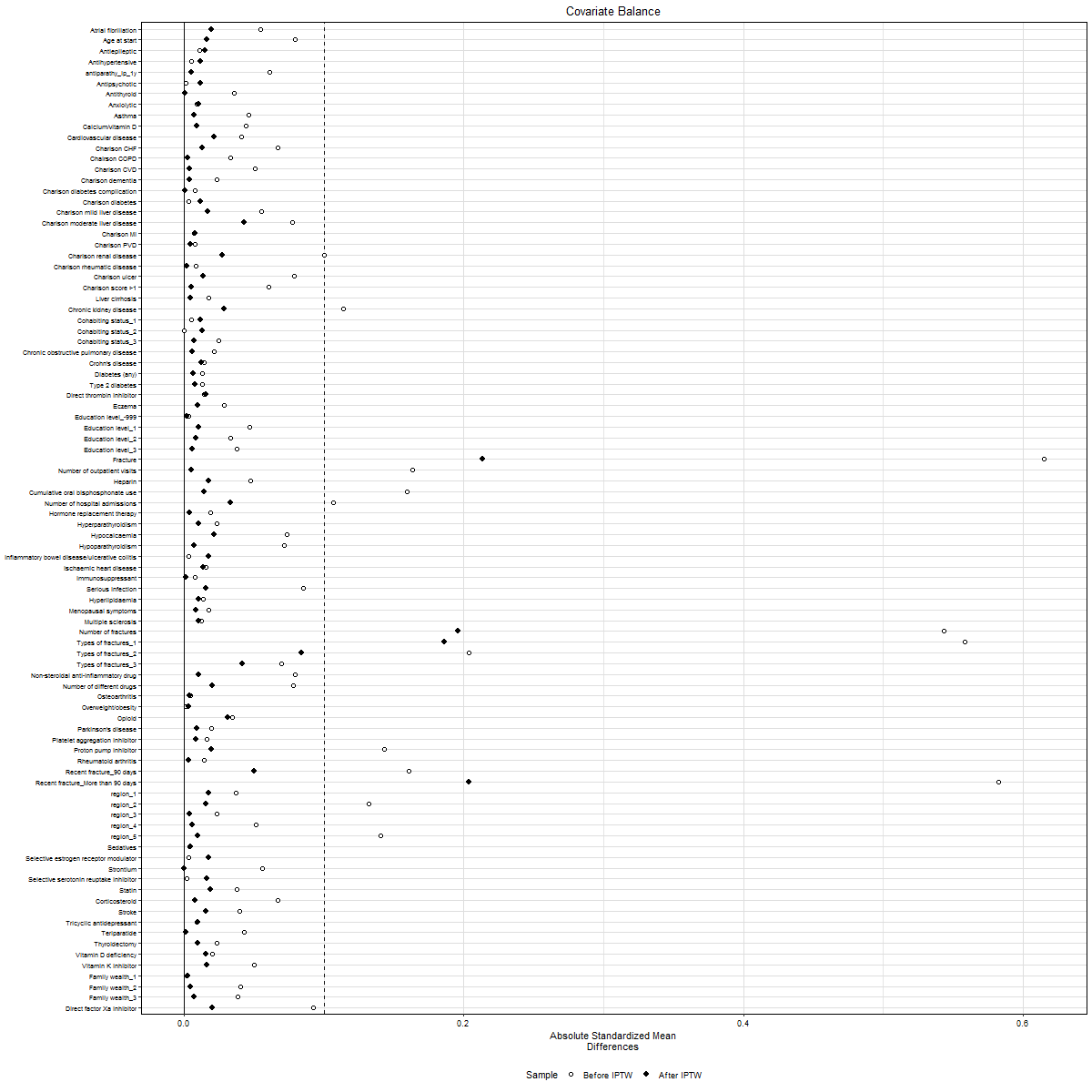

Table S2: Number of imbalanced covariates in new user cohort subgroups

|  | CPRD | DNR |
| --- | --- | --- |
| Subgroup | n/N, (%) of imbalanced covariates | n/N, (%) of imbalanced covariates |
| Older patients | 14/87, (17.1%) | 1/83, (1.2%) |
| Post-fracture patients | 15/87, (17.2%) | 2/83 (2.4%) |
| Patients with potentially 3 years of follow-up | 14/87 (17.1%) | 2/83 (2.4%) |

CPRD: Clinical Practice Research Datalink; DNR: Danish National Registries

N refers to the number of covariates in the PS model; n refers to the number of imbalanced covariates after PS methods

Note: IPTW was used for assessing covariate balance

Table S3: Comparability threshold using negative control outcome estimates in new user cohort subgroups

|  | **CPRD** | | **DNR** | |
| --- | --- | --- | --- | --- |
| **Subgroup** | **N** | **Approach 2**  **n (%)** | **N** | **Approach 2**  **n (%)** |
| Older patients | 41 | 2 (4.9) | 25 | 0 (0.0) |
| Post-fracture patients | 30 | 1 (3.3) | 12 | 0 (0.0) |
| Patients with potentially 3 years of follow-up | 43 | 2 (4.7) | 26 | 1 (3.8) |

CPRD: Clinical Practice Research Datalink; DNR: Danish National Registries

N refers to number of negative control outcomes with at least 100 events during follow-up period. Results presented in number of negative control outcomes with residual bias [n (percentage)]

Note: Approach 2 was used for comparability threshold for negative control outcome estimates
